# Biologic and clinical features of childhood gamma delta T-ALL: identification of STAG2/LMO2 γδ T-ALL as an extremely high risk leukemia in the very young

**DOI:** 10.1101/2023.11.06.23298028

**Authors:** Shunsuke Kimura, Petri Polonen, Lindsey Montefiori, Chun Shik Park, Ilaria Iacobucci, Allen EJ Yeoh, Andishe Attarbaschi, Andrew S. Moore, Anthony Brown, Atsushi Manabe, Barbara Buldini, Burgess B. Freeman, Chelsey Chen, Cheng Cheng, Chiew Kean Hui, Chi-Kong Li, Ching-Hon Pui, Chunxu Qu, Daisuke Tomizawa, David T. Teachey, Elena Varotto, Elisabeth M Paietta, Elizabeth D. Arnold, Franco Locatelli, Gabriele Escherich, Hannah Elisa Muhle, Hanne Vibeke Marquart, Hester A. de Groot-Kruseman, Jacob M. Rowe, Jan Stary, Jan Trka, John Kim Choi, Jules P.P. Meijerink, Jun J. Yang, Junko Takita, Katarzyna Pawinska-Wasikowska, Kathryn G. Roberts, Katie Han, Kenneth J. Caldwell, Kjeld Schmiegelow, Kristine R. Crews, Mariko Eguchi, Martin Schrappe, Martin Zimmerman, Masatoshi Takagi, Mellissa Maybury, Michael Svaton, Michaela Reiterova, Michal Kicinski, Mollie S. Prater, Motohiro Kato, Noemi Reyes, Orietta Spinelli, Paul Thomas, Pauline Mazilier, Qingsong Gao, Riccardo Masetti, Rishi S Kotecha, Rob Pieters, Sarah Elitzur, Selina M. Luger, Sharnise Mitchell, Shondra M. Pruett-Miller, Shuhong Shen, Sima Jeha, Stefan Köhrer, Steven M. Kornblau, Szymon Skoczeń, Takako Miyamura, Tiffaney L Vincent, Toshihiko Imamura, Valentino Conter, Yanjing Tang, Yen-Chun Liu, Yunchao Chang, Zhaohui Gu, Zhongshan Cheng, Zhou Yinmei, Hiroto Inaba, Charles G. Mullighan

## Abstract

**PURPOSE:** Gamma delta T-cell receptor-positive acute lymphoblastic leukemia (γδ T-ALL) is a high-risk but poorly characterized disease.

**METHODS:** We studied clinical features of 200 pediatric γδ T-ALL, and compared the prognosis of 93 cases to 1,067 protocol-matched non-γδ T-ALL. Genomic features were defined by transcriptome and genome sequencing. Experimental modeling was used to examine the mechanistic impacts of genomic alterations. Therapeutic vulnerabilities were identified by high throughput drug screening of cell lines and xenografts.

**RESULTS:** γδ T-ALL in children under three was extremely high-risk with 5-year event-free survival (33% v. 70% [age 3-<10] and 73% [age ≥10], *P*=9.5 x 10^-5^) and 5-year overall survival (49% v. 78% [age 3-<10] and 81% [age ≥10], *P*=0.002), differences not observed in non-γδ T-ALL. γδ T-ALL in this age group was enriched for genomic alterations activating *LMO2* activation and inactivating *STAG2* inactivation (*STAG2/LMO2*). Mechanistically, we show that inactivation of STAG2 profoundly perturbs chromatin organization by altering enhancer-promoter looping resulting in deregulation of gene expression associated with T-cell differentiation. Drug screening showed resistance to prednisolone, consistent with clinical slow treatment response, but identified a vulnerability in DNA repair pathways arising from STAG2 inactivation, which was efficaciously targeted by Poly(ADP-ribose) polymerase (PARP) inhibition, with synergism with HDAC inhibitors. Ex-vivo drug screening on PDX cells validated the efficacy of PARP inhibitors as well as other potential targets including nelarabine.

**CONCLUSION:** γδ T-ALL in children under the age of three is extremely high-risk and enriched for *STAG2/LMO2* ALL. STAG2 loss perturbs chromatin conformation and differentiation, and *STAG2/LMO2* ALL is sensitive to PARP inhibition. These data provide a diagnostic and therapeutic framework for pediatric γδ T-ALL.

**SUPPORT:** The authors are supported by the American and Lebanese Syrian Associated Charities of St Jude Children’s Research Hospital, NCI grants R35 CA197695, P50 CA021765 (C.G.M.), the Henry Schueler 41&9 Foundation (C.G.M.), and a St. Baldrick’s Foundation Robert J. Arceci Innovation Award (C.G.M.), Gabriella Miller Kids First X01HD100702 (D.T.T and C.G.M.) and R03CA256550 (D.T.T. and C.G.M.), F32 5F32CA254140 (L.M.), and a Garwood Postdoctoral Fellowship of the Hematological Malignancies Program of the St Jude Children’s Research Hospital Comprehensive Cancer Center (S.K.). This project was supported by the National Cancer Institute of the National Institutes of Health under the following award numbers: U10CA180820, UG1CA189859, U24CA114766, U10CA180899, U10CA180866 and U24CA196173.

**DISCLAIMER:** The content is solely the responsibility of the authors and does not necessarily represent the official views of the National Institutes of Health. The funding agencies were not directly involved in the design of the study, gathering, analysis and interpretation of the data, writing of the manuscript, or decision to submit the manuscript for publication.

## INTRODUCTION

The outcome of T-cell acute lymphoblastic leukemia (T-ALL) has improved with recent changes in therapeutic strategy including the incorporation of new drugs such as nelarabine as well as minimal residual disease (MRD)-based risk adaptation,^1–4^ however outcomes remain inferior to B-ALL. One group of high-risk T-ALL that is potentially associated with higher rates of refractory disease and poor prognosis is gamma delta (γδ) T-ALL, which expresses the γδ T-cell receptor (TCR) and accounts for 10% of T-ALL.^5,6^ Yet, the clinical features and prognostic significance of γδ T-ALL have been only examined in small cohorts.^5,6^

Genomic analyses have identified multiple new subtypes and drivers of B-ALL, and have transformed the approach to diagnosis, risk classification, and for some subtypes, therapeutic targeting.^7–9^ Recent studies have examined the genomic landscape of T-ALL using whole exome sequencing (WES) and whole transcriptome sequencing (RNAseq), and have defined several T-ALL subtypes arising from dysregulated oncogene expression.^10,11^ However, these studies included few documented cases of γδ T-ALL, and the genomic basis of this form of leukemia remains elusive. Moreover, these genomic studies have demonstrated the limitations of non-whole genome sequencing (WGS) studies of T-ALL, in which oncogene-deregulating genomic alterations commonly involve non-coding regions of the genome that are not detected by RNAseq or WES, and require WGS and chromatin profiling approaches such as HiChIP to fully elucidate driver genomic alterations.^12,13^

Thus, limited studies have explored the clinical and genomic characteristics of γδ T-ALL. In this study, we sought to conduct a comprehensive analysis of this entity, to identify outcome determinants, and to explore the therapeutic potential for better classification and improved outcomes.

## METHODS

### Study Design

Data from all patients with T-ALL expressing γδ TCR up to 25 years of age enrolled in clinical trials and diagnosed from January 1, 2000, through December 31, 2018, were retrospectively collected from 13 cooperative study groups (n=200, **Appendix Table A1**). Data from patients with non-γδ T-ALL (n=1,067) enrolled on the same protocols (DCOG-ALL10,^14^ AIEOP-BFM ALL2000,^4^ EORTC 58951,^15^ SJCRH-T15/T16^16,17^) were collected for comparison analysis (**Fig 1A**). Only the protocol-matched patients from these four groups were used for the outcome analyses; for the analyses of clinical features, we used all 200 γδ T-ALL data. All clinical trials were approved by institutional review boards or ethics committees. This study was approved by the Institutional Review Board of St. Jude Children’s Research Hospital.

**Figure 1.**
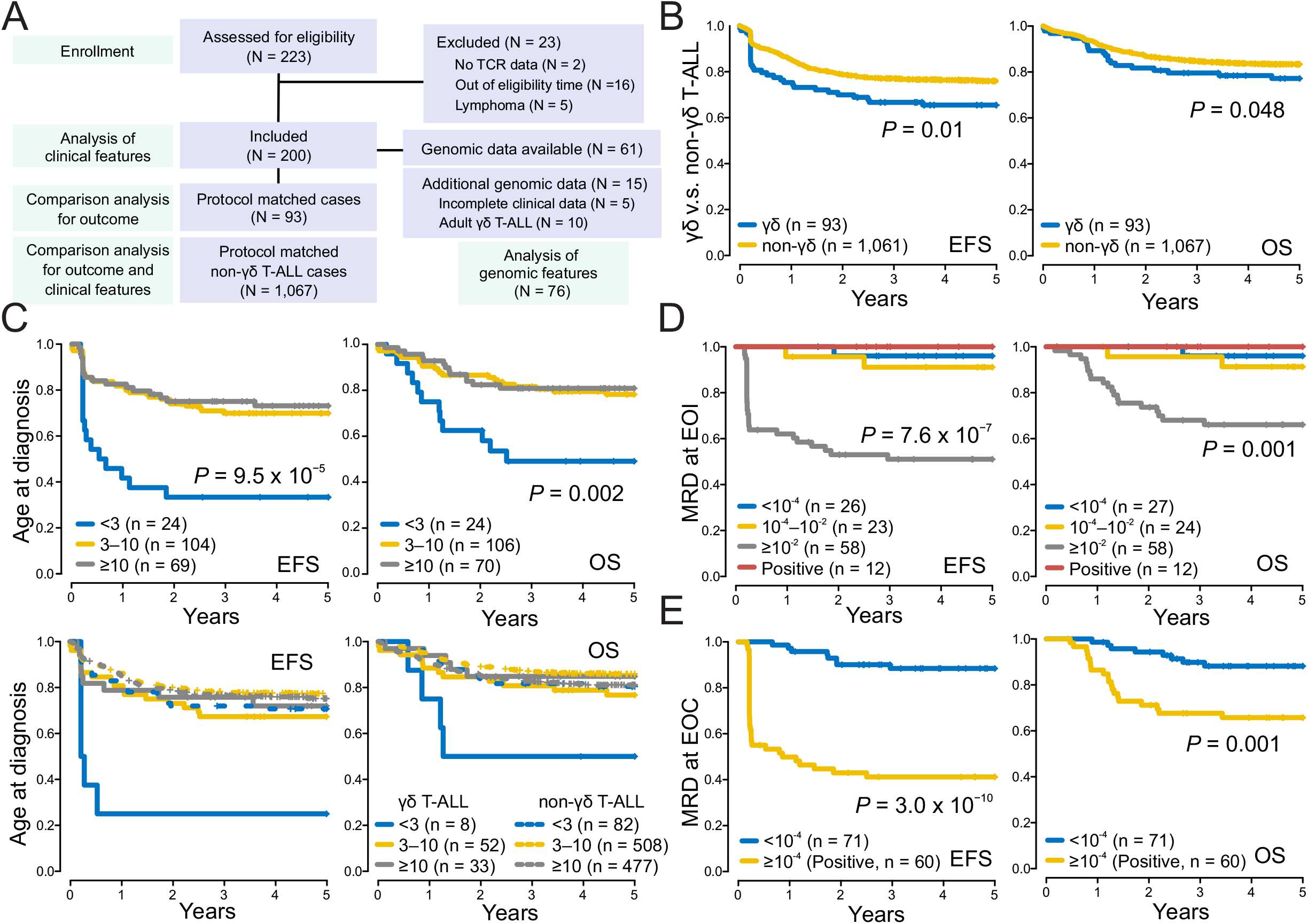
Study cohort and outcome of γδ and non-γδ T-ALL. **A,** CONSORT diagram. **B,** γδ T-ALL showed significantly worse overall survival (OS) and event-free survival (EFS) compared to non-γδ T-ALL. **C,** Poor OS and EFS of γδ T-ALL for children under three years of age (top panels); outcomes of non-γδ T-ALL (dotted line) did not vary by age (bottom panels). **D,** γδ T-ALL patients with MRD≥10^-2^ (1%) at EOI exhibited significantly worse OS and EFS than those with MRD<10^-2^. **E,** γδ T-ALL patients with positive MRD (≥10^-4^) at the end of consolidation (EOC) had inferior outcomes to those with undetectable MRD. The *P* values were calculated using the log-rank test.

### Terminology and Outcomes

The following definitions were used to analyze clinical features according to an international consensus of the Ponte-di-Legno Consortium.^18^ The definition of the time points at the end of induction (EOI) and consolidation (EOC) was approximately four weeks and 7-12 weeks (day 42/46 for SJCRH-T15/T16) from the start of treatment, respectively (**Appendix Table A1**). Complete remission (CR) was defined when bone marrow (BM) showed M1 cytomorphology and/or MRD <1% without evidence of extramedullary disease at EOC. When MRD data were available, MRD results were preferentially used to define CR. MRD was measured by PCR and/or flow cytometry depending on the study. Failure to achieve CR was defined as a treatment failure event on EOC assessment day. Event-free survival (EFS) was defined as the time from the start of treatment to any of the first events including the death of any time, treatment failure, relapse, second malignancy, or last follow-up date for those who were event-free. Overall survival (OS) was defined as the time from the start of treatment to death from any cause or last follow-up date.

### Patient samples, sequencing, gene-edited models, and drug screening

Details for the process of primary materials, WGS, RNAseq, HiChIP, chromatin immunoprecipitation sequencing (ChIP-seq), and methods for cell culture, generation of CRISPR/Cas9-based gene-edited models, and drug screening are described in the Data Supplement, **Supplementary Methods**.

### Statistical Analysis

Statistical analyses were performed using R v.4.2.2 software. Categorical variables were analyzed using the Chi-square test. The two-sided Wilcoxon rank-sum test was used to compare the mean values of different groups. Two-sided log-rank tests were used for survival analyses. For univariate and multivariate analyses of prognostic factors, the Cox proportional hazards regression model was used.

## RESULTS

### Identification of a subset of high-risk **γδ** T-ALL in the very young

A total of 200 cases of γδ T-ALL patients were assembled through a consortium of 13 ALL study groups (**Appendix Table A1**). From the analysis of those treated in the same clinical trials, the frequency of γδ T-ALL was 8.0% of T-ALL (n=93 and 1,067 for γδ T-ALL and non-γδ T-ALL, respectively, **Fig 1A**), and γδ T-ALL patients exhibited significantly inferior 5-year EFS (65% v. 76%, *P*=0.01) and OS (77% v. 83%, *P*=0.048) compared to non-γδ T-ALL patients (**Fig 1B**). In comparison with non-γδ T-ALL, γδ T-ALL patients were characterized by younger age at diagnosis (*P*=0.01), higher white blood cell (WBC) at diagnosis (*P*=0.02), less frequent mediastinal mass (*P*=9.1×10^-5^), and a more common mature T immunophenotype (surface (s)CD3+, CD1a-, *P*=1.3×10^-15^, **Table 1**, and **Appendix Table A2**). MRD measurements were available for 90, 121, and 131 patients at day 15, EOI, and EOC, respectively. γδ T-ALL patients exhibited a higher rate of poor prednisone response (*P*=0.0008), MRD ≥1% (≥10^-2^) at day 15 (*P*=4.3 x 10^-7^), at EOI (*P*=6.4 x 10^-8^) and at EOC (*P*=6.1 x 10^-8^, **Table 1**, and **Appendix Table A2**), suggesting slow treatment response and chemoresistance leading to treatment failure. Accordingly, more γδ T-ALL patients underwent hematopoietic stem cell transplantation (*P*=6.4 x 10^-8^) and died from primary disease (16.8% v. 11.2%) and toxicity during treatment compared to non-γδ T-ALL patients (7.6% v. 4.0%, *P*=0.0004, **Table 1**, and **Appendix Table A2**). The cumulative incidence of relapse (**Appendix Fig A1A**) and OS after relapse and treatment failure in γδ T-ALL were similar to that of non-γδ T-ALL (**Appendix Fig A1B,C**). Importantly, there was a significant difference in the site of relapse; almost all relapses of γδ T-ALL were isolated BM relapses (91.3% v. 50.0%), while central nervous system (CNS, isolated CNS and combined CNS/BM) was more commonly involved in non-γδ T-ALL (4.3% v. 41.6%, *P* = 0.003, **Table 1**, and **Appendix Fig A1D**). No difference was observed in CNS involvement at diagnosis between γδ and non-γδ T-ALL (**Appendix Table A2**).

**Table 1.** Comparison of clinical characteristics and outcomes between gamma delta T-ALL and non-gamma delta T-ALL.

| Characteristic | No. of $\gamma\delta$ T-ALL Patients | No. of non-$\gamma\delta$ T-ALL Patients | Chi square test | $\gamma\delta$ T-ALL | | | | | |
| --- | --- | --- | --- | --- | --- | --- | --- | --- | --- |
|  |  |  |  | No. of Events | Five-Year event-Free Survival Rate, % (95% CI) | Log Rank P | No. of Deaths | Five-Year Overall Survival Rate, % (95% CI) | Log Rank P |
| Sex [n (%)] |  |  | 0.053 |  |  | 0.49 |  |  | 0.55 |
| Female | 70 (35%) | 301 (28.2%) |  | 26 | 63 (53–76) |  | 16 | 80 (71–90) |  |
| Male | 130 (65%) | 766 (71.8%) |  | 42 | 68 (61–77) |  | 34 | 73 (66–82) |  |
| Age at Dx [n (%)] | | | 0.01 | | | $9.49 \times 10^{-5}$ | | | 0.002 |
| <3 | 24 (12%) | 82 (7.7%) |  | 16 | 33 (19–59) |  | 12 | 49 (33–74) |  |
| 3 - <10 | 106 (53%) | 508 (47.6%) |  | 33 | 70 (62–79) |  | 23 | 78 (71–87) |  |
| ≥10 | 70 (35%) | 477 (44.7%) |  | 19 | 73 (63–85) |  | 15 | 81 (72–91) |  |
| WBC at DX [n (%)] |  |  | 0.02 |  |  | 0.74 |  |  | 0.73 |
| 0 - <20 | 44 (22%) | 335 (31.4%) |  | 13 | 72 (59–87) |  | 9 | 78 (67–92) |  |
| 20 - <50 | 36 (18%) | 169 (15.8%) |  | 13 | 63 (49–81) |  | 10 | 72 (57–90) |  |
| ≥50 | 120 (60%) | 556 (52.1%) |  | 42 | 66 (58–75) |  | 31 | 75 (68–84) |  |
| NA | 0 | 7 |  |  |  |  |  |  |  |
| PSL Response [n (%)] |  |  | 0.0008 |  |  | 0.0002 |  |  | 0.02 |
| Responder | 78 (52.3%) | 402 (67%) |  | 17 | 79 (71–89) |  | 13 | 83 (75–92) |  |
| Non-Responder | 71 (47.7%) | 198 (33%) |  | 34 | 52 (41–65) |  | 23 | 69 (59–81) |  |
| NA | 51 | 467 |  |  |  |  |  |  |  |
| MRD at day15 [n (%)] | | | $4.34 \times 10^{-7}$ | | | 0.002 | | | 0.03 |
| <10<sup>-4</sup> | 5 (5.5%) | 36 (26.3%) |  | 1 | 80 (52–100) |  | 1 | 80 (52–100) |  |
| 10<sup>-4</sup> – 10<sup>-2</sup> | 18 (19.8%) | 47 (34.3%) |  | 0 | 100 (100–100) |  | 0 | 100 (100–100) |  |
| ≥10<sup>-2</sup> | 67 (74.7%) | 54 (39.4%) |  | 31 | 52 (41–65) |  | 21 | 67 (56–80) |  |
| NA | 110 | 930 |  |  |  |  |  |  |  |
| MRD at EOI [n (%)] | | | $6.41 \times 10^{-8}$ | | | $7.64 \times 10^{-7}$ | | | 0.001 |
| <10<sup>-4</sup> | 27 (24.8%) | 134 (27.0%) |  | 1 | 96 (89–100) |  | 1 | 96 (89–100) |  |
| 10<sup>-4</sup> – 10<sup>-2</sup> | 24 (22.0%) | 231 (46.5%) |  | 3 | 91 (80–100) |  | 3 | 91 (80–100) |  |
| ≥10<sup>-2</sup> | 58 (58.0%) | 132 (26.6%) |  | 29 | 51 (40–66) |  | 20 | 66 (55–80) |  |
| Positive | 12 | 30 |  | 0 | 100 (100–100) |  | 0 | 100 (100–100) |  |
| NA | 79 | 540 |  |  |  |  |  |  |  |
| MRD at EOC [n (%)] | | | 0.02 | | | $3.00 \times 10^{-10}$ | | | 0.001 |
| <10<sup>-4</sup> | 71 (54.2%) | 424 (64.6%) |  | 9 | 88 (81–96) |  | 9 | 88 (81–96) |  |
| Positive | 60 (45.8%) | 232 (35.4%) |  | 36 | 41 (30–56) |  | 21 | 66 (55–79) |  |
| NA | 69 | 411 |  |  |  |  |  |  |  |
| Total relapses | 23 | 127 | 0.003 |  |  |  |  |  |  |
| BM | 21 (91.3%) | 60 (50.0%) |  |  |  |  |  |  |  |
| BM combined | 1 (4.3%) | 19 (15.8%) |  |  |  |  |  |  |  |
| CNS | 0 | 31 (25.8%) |  |  |  |  |  |  |  |
| Others | 1 (4.3%) | 10 (8.3%) |  |  |  |  |  |  |  |
| Unknown | 0 | 7 |  |  |  |  |  |  |  |
| Death reasons |  |  | 0.0004 |  |  |  |  |  |  |
| Alive cases | 150 | 871 |  |  |  |  |  |  |  |
| Total death cases | 50 | 191 |  |  |  |  |  |  |  |
| ALL | 29 (16.8%) | 54 (11.2%) |  |  |  |  |  |  |  |
| Toxicity | 13 (7.6%) | 19 (4.0%) |  |  |  |  |  |  |  |
| Second malignancy | 1 (0.6%) | 7 (1.5%) |  |  |  |  |  |  |  |
| Unknown | 7 | 111 |  |  |  |  |  |  |  |

Strikingly, γδ T-ALL diagnosed before three years of age exhibited markedly inferior EFS compared to those diagnosed in older children (33% v. 70% [age 3-<10] and 73% [age ≥10], *P*=9.5 x 10^-5^) and OS (49% v. 78% [age 3-<10] and 81% [age ≥10], *P*=0.002), a difference that was not observed in non-γδ T-ALL (EFS; 72% v. 80% [age 3-<10] and 76% [age ≥10], OS; 80% v. 86% [age 3-<10] and 81% [age ≥10]) (**Fig 1C**, **Table 1**, and **Appendix Table A2**). Eleven out of 24 γδ T-ALL patients under three years old died from primary disease or toxicity. In addition, γδ T-ALL with MRD ≥1% at EOI showed poor EFS (51% v. 96% [below 0.01%] and 91% [0.01%–1%], *P*=7.6×10^-7^) and most were not salvaged (OS, 66% v. 96% [below 0.01%] and 91% [0.01%–1%], *P*=0.001, **Fig 1D** and **Table 1**). Positivity (≥0.01%) of MRD at EOC also predicted poor EFS (41% v. 88%, *P*=3.0×10^-10^) and OS (66% v. 88%, *P*=0.001, **Fig 1E**, **Table 1**, and **Appendix Fig A1E**). Forty-four out of 53 (83%) patients with MRD ≥1% at EOI remained MRD positive at EOC and 34% (n=18) of them died (**Appendix Fig A1F**). In a multivariable analysis, age under three was associated with worse EFS and OS, though MRD levels at EOI (≥1%) and EOC (≥0.01%) remained important prognostic factors in MRD-directed protocols (**Appendix Table A3**). The use of MRD-stratified protocols did not improve the outcome of γδ T-ALL (**Appendix Table A2** and **Appendix Fig A1G**). Thus, clinical data identified two high-risk groups in γδ T-ALL - those under three years of age and those with MRD ≥1% at EOI – that required alternative therapy, rather than intensification with conventional treatment.

### Enrichment of the *STAG2/LMO2* subtype in high-risk **γδ** T-ALL in the young

To determine the genetic basis of γδ T-ALL, WGS (n=47) and RNAseq (n=68) were performed, including 15 additional γδ T-ALL samples (**Appendix Tables A4–A7**). We combined analysis of these samples with 1,076 T-ALL cases whose TCR status at diagnosis was not available (Children’s Oncology Group AALL0434 study^2,19^). Gene expression profiling revealed that γδ T-ALL cases were classified by leukemia-initiating events and maturation stages (**Fig 2A**, **Appendix Fig A2,** and **Appendix Table A8**). γδ T-ALL was enriched for several subtypes including *LMO2* γδ-like, T-ALL with recurrent chromosome gains (Chr gains), *STAG2/LMO2*, *TLX3*-rearrangement, and *PICALM::MLLT10* (**Fig 2B**). Each of these subtypes except for *STAG2/LMO2* is described in Data Supplement, **Supplementary Results, Appendix Fig A3,4,** and **Appendix Table A9**.

**Figure 2.**
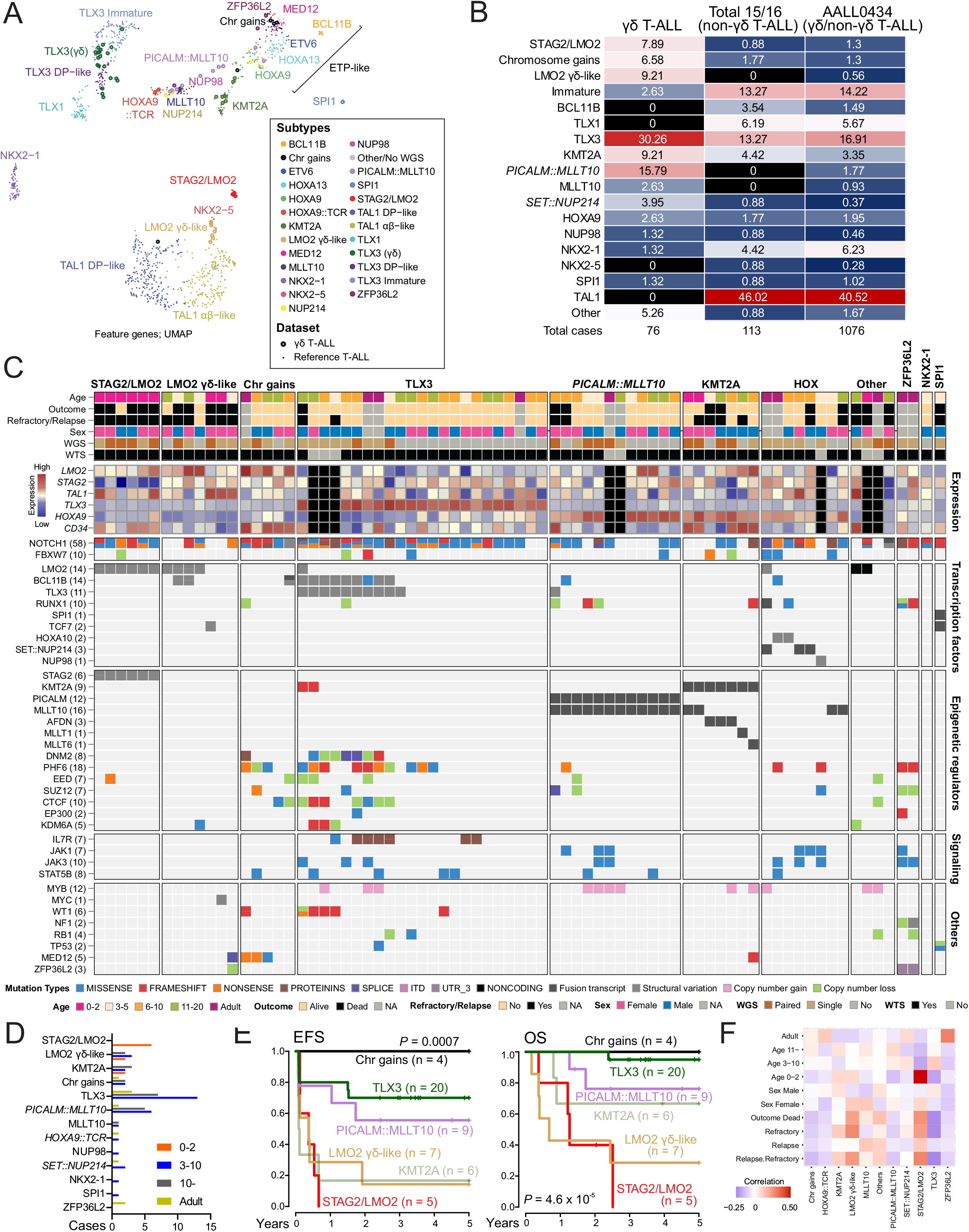
The intersection of genomics and clinical features of γδ T-ALL. **A,** UMAP plot of gene expression analysis for 68 cases of γδ T-ALL (large bold circles) layered on the reference T-ALL cohort (n=1,076, small circles)^19^. **B,** Frequency of each genomic subtype in the γδ T-ALL cohort (n=76), non-γδ T-ALL cases from Total 15/16 of St. Jude Children’s Research Hospital (n=113), and AALL0434 cohorts (no TCR data at diagnosis was available, n=1,076). **C,** Heatmap showing the mutational landscape, the expression level of selected subtype defining genes, and clinical parameters of the 76 cases of γδ T-ALL. Black color in expression indicates no data. **D,** Age distribution of each genomic subtype in γδ T-ALL cases (n = 76). **E,** Event-free survival (EFS) and overall survival (OS) in each genomic subtype. The *P* value was calculated using the log-rank test. **F,** Correlation of clinical features and genomic subtypes.

*STAG2/LMO2* is a subtype enriched in γδ T-ALL that has a distinct expression profile (7.9% for γδ T-ALL v. 0.9% in non-γδ T-ALL, **Fig 2A,B**). All *STAG2/LMO2* cases had dual-hit alterations targeting *LMO2* and *STAG2* (**Fig 2C**). All *STAG2/LMO2* γδ cases were diagnosed before the age of three, including infancy, a pattern of age at diagnosis not observed in other γδ T-ALL subtypes (**Fig 2D**).

The clinical outcomes of γδ T-ALL exhibited significant variation depending on the genomic subtype. The Chr gains and *TLX3*-rearranged subtypes had excellent OS (**Fig 2E**). In contrast, the *STAG2/LMO2*, *LMO2* γδ-like, and *KMT2A*-rearranged subtypes had poor EFS, and notably, these subtypes included all γδ T-ALL patients diagnosed under the age of three (**Fig 2D,E**). Among these three γδ T-ALL subtypes, *STAG2/LMO2* was predominantly related to young onset under three, indicating the overlap of high-risk groups in γδ T-ALL (**Fig 2F**). Indeed, three out of five *STAG2/LMO2* cases succumbed to death by the primary disease. Thus, integrated clinico-genomic analyses demonstrated that the *STAG2/LMO2* subtype is enriched in children diagnosed with T-ALL under the age of three and the γδ immunophenotype, who had dismal outcomes.

### Dual hit genomic drivers in *STAG2/LMO2* T-ALL

To further elucidate the biology of *STAG2/LMO2* subtype, we collected and analyzed 18 additional *STAG2/LMO2* cases including non-γδ T-ALL (total 24 cases, **Appendix Table A10**). All but 2 (both from additional cases and diagnosed before 4 years of age) were diagnosed before age three and included five infant T-ALL cases (**Fig 3A**). Unlike the general male sex predominance in T-ALL, female sex was more common (58%) in *STAG2/LMO2* T-ALL.

**Figure 3.**
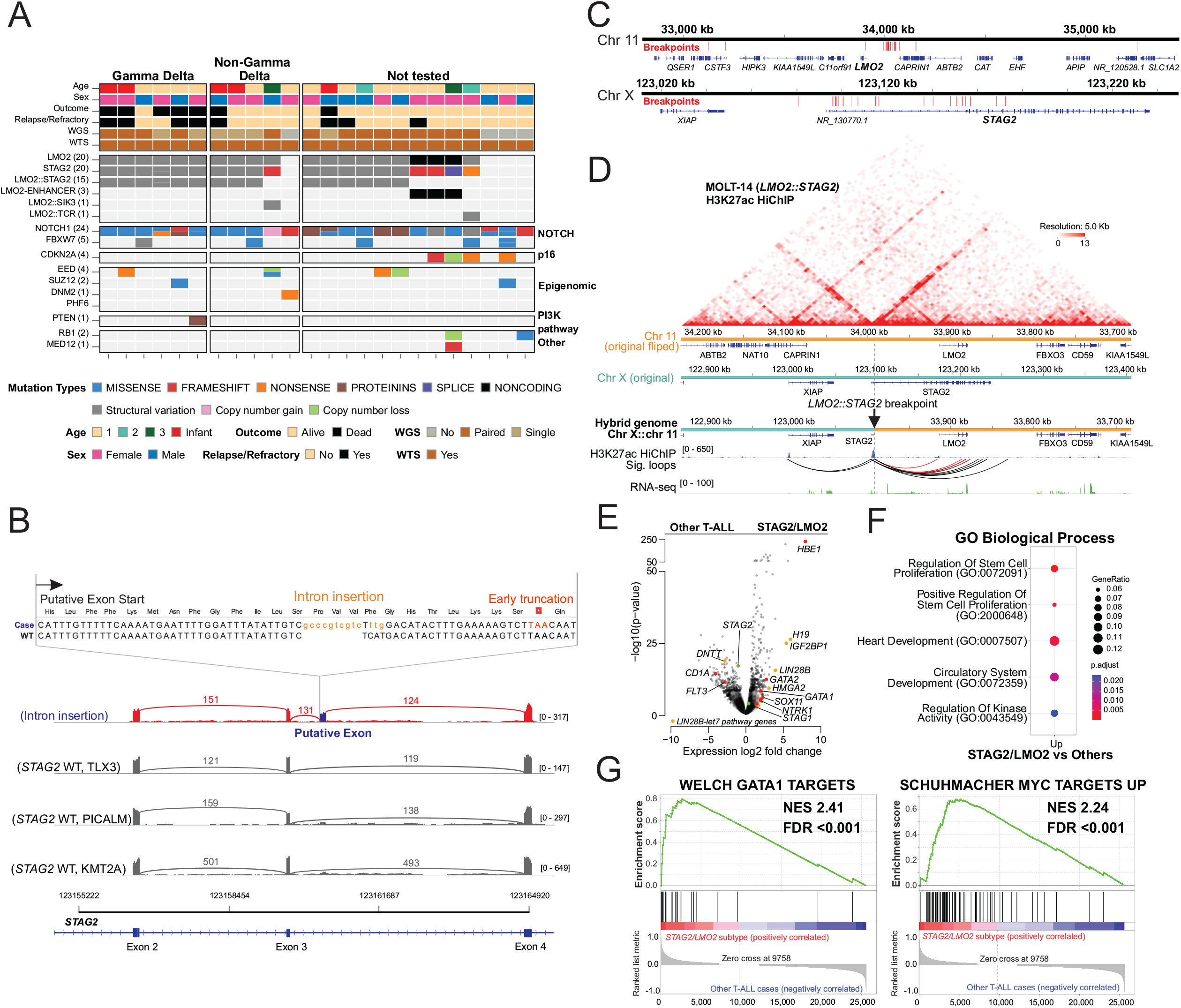
The genomics of *STAG2/LMO2* T-ALL. **A,** Heatmap showing the mutational landscape and clinical parameters of the 24 cases of *STAG2/LMO2* T-ALL, including 6 cases of γδ T-ALL, 5 of non-γδ T-ALL, and 13 lacking TCR status at diagnosis (n=13). **B,** Example of variant alterations of STAG2 inactivation. One case harbored a 13 bp indel alteration in intron 3 of *STAG2*, resulting in a putative exon, aberrant splicing and early STAG2 truncation. Sashimi plots shows the splicing between *STAG2* exon 2 and exon 4. Intron indel alterations (orange) deregulate splicing of this region, generating a putative exon between exon 3 and 4 with early truncation. **C,** Breakpoints of *LMO2::STAG2* translocation are shown in red bar. **D,** H3K27ac HiChIP on MOLT-14 cell line with *LMO2::STAG2* is shown. Data was aligned on a custom reference that mimics chromosome 11 (orange) and X (cyan) translocation in MOLT-14. The top heat map represents raw interaction, and HiChIP and RNAseq coverage tracks are shown in the middle and the bottom, respectively. Significant H3K27ac-anchored interactions (FDR<0.01) are shown as arcs. Interactions between the *STAG2* promoter and the *LMO2* gene are shown in red. FDR, false discovery rate. **E,** Differentially expressed genes in *STAG2/LMO2* cases (n=24) compared with other T-ALL (n=1,120) are shown in the volcano plot. LIN28/let-7 pathway genes are colored orange. Cohesin complex genes (*STAG1* and *STAG2*) are colored green. **F,** Pathway analysis (GO Biological Process) using up-regulated genes (n=78, adjusted *P*<0.01 and fold change >2) in *STAG2/LMO2* compared to other T-ALL cases. **G,** Up-regulated pathways in *STAG2/LMO2* subtype compared to other T-ALL cases analyzed by gene set enrichment analysis.

Dual-hit alterations at the *LMO2* and *STAG2* loci were identified in all 20 cases with WGS data. Most commonly, these included *LMO2::STAG2* rearrangements (n=15, 75%); alternatively, *LMO2* enhancer single nucleotide variants or rearrangements to TCR loci that resulted in aberrant *LMO2* expression, were present with concurrent *STAG2* alterations leading to premature truncation (**Fig 3A,B** and **Appendix Table A10**). *LMO2::STAG2* does not generate a fusion chimera, thus these rearrangements could not be identified in cases lacking WGS. In most cases, breakpoints were detected upstream of *LMO2* and within the introns of *STAG2* (**Fig 3C**, Data supplement, **Supplementary Fig S1,** and **Appendix Table A10**). H3K27ac HiChIP to integrate enhancer profiling with chromatin looping was performed in the *STAG2/LMO2* MOLT-14 cell line. This showed abnormal looping between *STAG2* promoter and *LMO2* promoter/enhancers, indicating the induction of *LMO2* expression at the cost of *STAG2* expression (**Fig 3D** and **Appendix Fig A5A,B**). All cases had *NOTCH1* activating alterations, but few additional alterations, suggesting that *LMO2* deregulation, STAG2 inactivation, and NOTCH1 pathway activation are necessary and sufficient drivers of leukemogenesis.

*STAG2/LMO2* cases exhibited up-regulation of LIN28/let-7 pathway genes, which are expressed in fetal hematopoietic stem cells (HSCs) and silenced after birth and following HSC differentiation^20^ (**Fig 3E**, **Appendix Fig A5C–E**, and **Appendix Table A11**). *HBE1*, *GATA1* and *GATA2*, key genes for early-stage hematopoiesis, were also highly expressed (**Fig 3E**), suggesting a fetal HSC origin of *STAG2/LMO2* ALL. Pathway analysis using the up-regulated genes in *STAG2/LMO2* revealed the enrichment of pathways related to stem cell proliferation (**Fig 3F**). Furthermore, gene-set enrichment analysis revealed up-regulation of MYC and GATA1 target pathways (**Fig 3G** and **Appendix Table A12**).

### STAG2 inactivation in T-ALL induces epigenomic deregulation

Although dual-hit alterations of *LMO2* and *STAG2* are unique features of the *STAG2/LMO2* subtype, aberrant expression and/or alterations of *LMO2* alterations are frequently observed in other subtypes of T-ALL, especially those with deregulation of *TAL1*. However, *STAG2* alterations are specific for *STAG2/LMO2* T-ALL, and STAG2 inactivation may drive the pathogenesis and gene expression profiles of this subtype. STAG2 is a member of the cohesin complex which has a critical role in the maintenance of enhancer-promoter short loops.^21^ Therefore, loss of STAG2 alters CTCF-anchored loop extrusion and rewires enhancer-promoter loops.^22–24^ Accordingly, chromatin loop size defined by H3K27ac HiChIP (generally representing enhancer-promoter loops) was highest in *STAG2/LMO2* cell lines compared to normal thymocytes and T-ALL cases, with the exception of DND-41, a *TLX3*-rearranged T-ALL cell line with *CTCF* alteration (**Fig 4A**).

**Figure 4.**
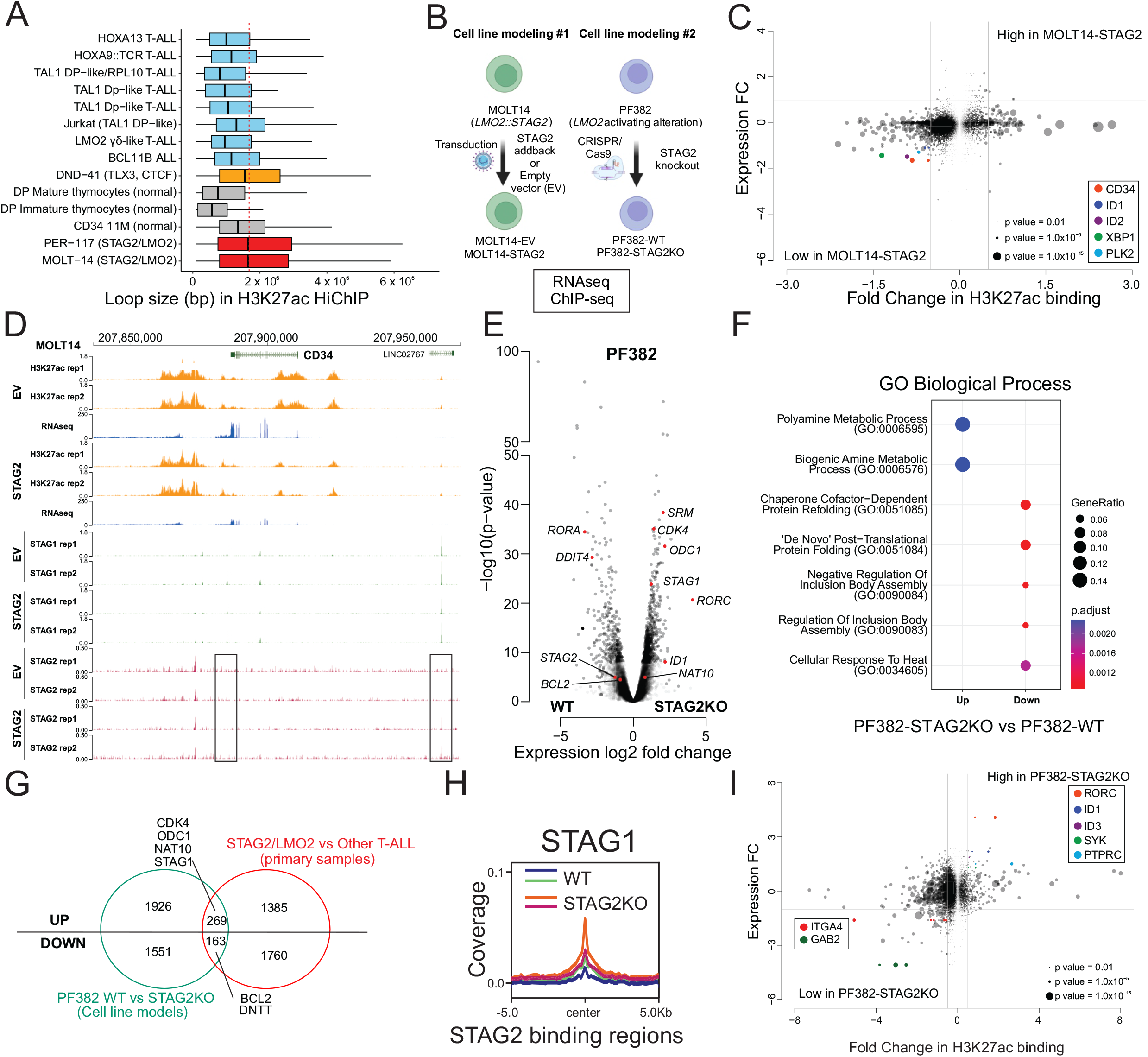
Effects of STAG2 inactivation in T-ALL cell line models. **A,** Chromatin loop size defined by H3K27ac HiChIP for representative T-ALL subtypes (blue and orange) and normal thymocytes (in grey). *STAG2/LMO2* cell lines are colored red. **B,** The schema of gene-edited cell line models. **C,** Starburst plot comparing MOLT14-EV with MOLT14-STAG2 for peaks of H3K27ac and their corresponding gene expression. Each circle indicates detected H3K27ac peaks by MACS2 and the circle size represents the p-value of each peak. The bottom-left section includes higher peaks with higher expression in MOLT14-EV. **D,** H3K27ac (orange), STAG1 (green), and STAG2 (red) binding and RNAseq (blue) coverage at the *CD34* locus in MOLT14-EV and MOLT14-STAG2 cells in duplicates. The black squares indicate regained STAG2 binding in MOLT14-STAG2. **E,** Differentially expressed genes in PF382-STAG2KO (n = 3) compared with PF382-EV (n = 2) in the volcano plot. **F,** Pathway analysis using up- or down-regulated genes (adjusted P <0.01 and fold change >2 or <-2) in PF382-STAG2KO compared to PF382-WT. **G,** Up- and down-regulated genes in PF382-STAG2 KO model (green) and primary *STAG2/LMO2* subtype T-ALL cases (red) are shown. **H,** Average ChIP-seq coverage of STAG1 in PF382-WT and PF382-STAG2 KO lines around common STAG1/STAG2, STAG1, and STAG2 binding regions detected from PF382-WT. **I,** Starburst plot comparing PF382-STAG2 KO with PF382-WT for peaks of H3K27ac and their corresponding gene expression. The top-right section indicates higher peaks with higher expression in PF382-STAG2 KO.

To further explore the effect of STAG2 inactivation in T-ALL, we developed two gene-edited cell line models: (1) empty vector (EV) or *STAG2* transduced MOLT-14 (*STAG2/LMO2*), and (2) *STAG2* knockout PF382 (*LMO2*-activating mutation; **Fig 4B**, Data supplement, **Supplementary Results,** and **Appendix Fig A6,A7**). Comparison of MOLT14-EV (STAG2 inactivation) and MOLT14-STAG2 (STAG2 restoration) in H3K27ac binding peaks and corresponding gene expression levels revealed genes regulated by STAG2 expression, including *CD34*, *ID1* and *ID2* (**Fig 4C,D** and **Appendix Table A13**). Pathway analysis (**Appendix Fig A6D**) suggested differentiation arrest induced by STAG2 inactivation.

We next examined the effects of STAG2 inactivation by using PF382 parental and *STAG2* knockout lines. STAG2 inactivation resulted in de-regulation of genes associated with T-cell differentiation, cell cycle, and polyamine metabolism in addition to *STAG1* (**Fig 4E,F, Appendix Fig A6E** and **Appendix Table A14**). Comparison of differentially expressed genes between PF382 *STAG2* knockout *v.* parental cells, and *STAG2/LMO2* T-ALL *v.* other T-ALL cases revealed 269 up-regulated and 163 down-regulated genes in common (**Fig 4G, Appendix Fig A6F** and **Appendix Table A15**). STAG2 binding in PF382 cells was partly compensated by STAG1 after STAG2 inactivation, consistent with up-regulation of *STAG1* expression following STAG2 loss (**Fig 4H** and **Appendix Fig A6H,A7**). Differential peaks of H3K27 acetylation and corresponding gene expression changes between PF382 parental and *STAG2* knockout cells included T-cell differentiation-related genes as seen in the MOLT-14 model (**Fig 4I** and **Appendix Table A16**). Thus, STAG2 inactivation drives perturbation of gene expression in *STAG2/LMO2* T-ALL, and T-cell differentiation arrest by deregulating chromatin conformation.

### Therapeutic targeting of *STAG2/LMO2* T-ALL

To identify potential new therapeutic approaches for *STAG2/LMO2* T-ALL, we performed drug-response screening for 2,050 compounds in two *LMO2::STAG2* T-ALL cell lines: MOLT-14 (γδ T-ALL) and PER-117 (non-γδ T-ALL; **Appendix Table A17**). 138 compounds were active in both cell lines at 10 µM single-dose screening, and in subsequent dose-response testing, 85 exhibited moderate to high efficacy (EC_50_ <1 μM) in both cell lines (**Fig 5A** and **Appendix Table A18**). Multiple histone deacetylase (HDAC), topoisomerase, and bromodomain (BRD) inhibitors showed efficacy (**Appendix Fig A8A**). Notably, inhibitors of the deregulated *LIN28B* and *NTRK1* genes in the *STAG2/LMO2* subtype (**Fig 3E**), were ineffective (**Appendix Fig A8B–E**), suggesting that deregulated expression of these genes is reflective of cell stage or state but is not oncogenic or a therapeutic vulnerability.

**Figure 5.**
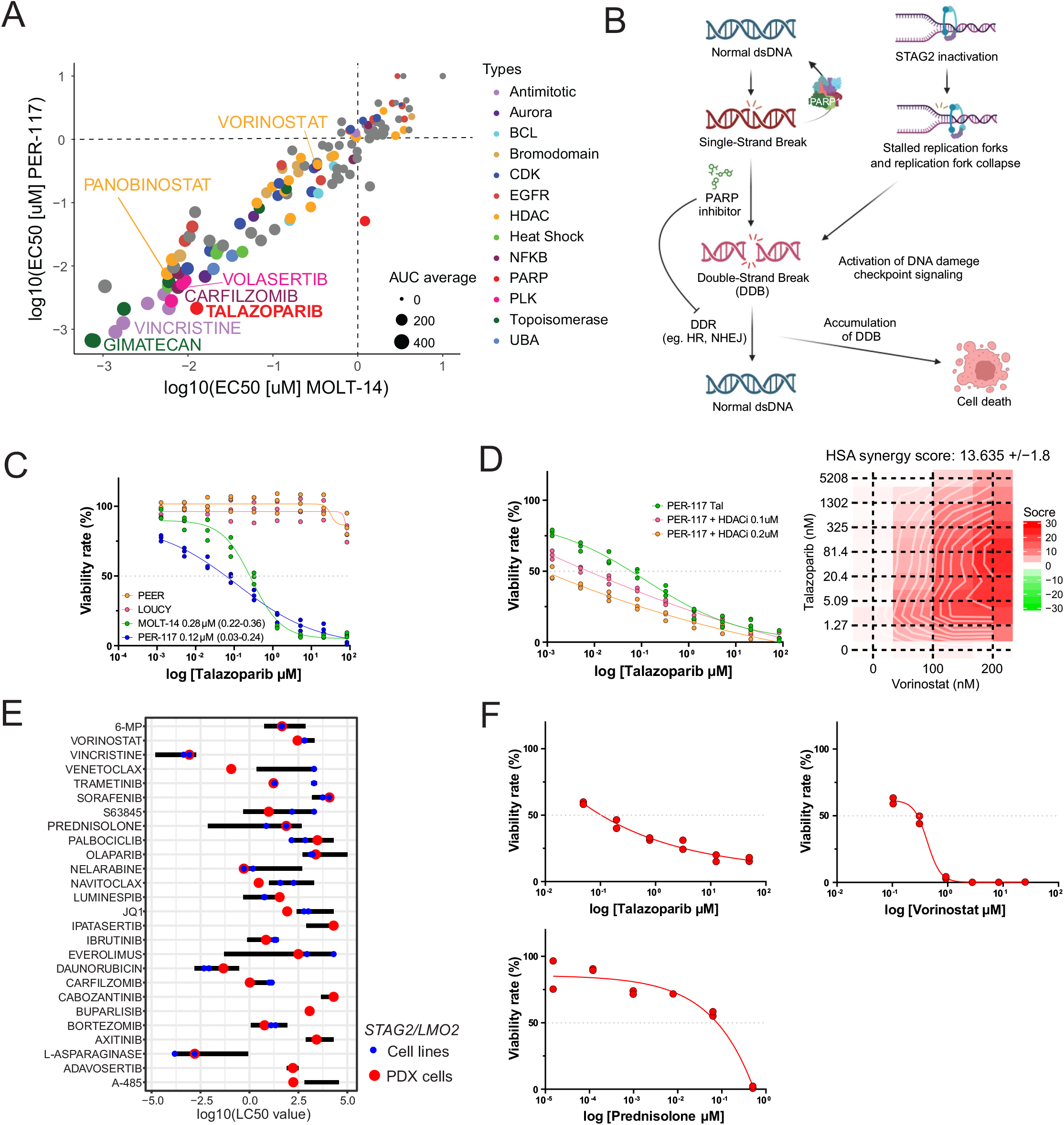
Drug screening in *STAG2/LMO2* T-ALL showed talazoparib, HDAC, and CDK inhibitors as potential target therapy. **A,** The result of the dose-response analysis for 138 compounds. Each circle indicates tested compounds and the circle size represents the average area under the curve (AUC). EC_50_ was calculated after 72 hours of treatment. EC_50_ <1 μM in both cell lines (bottom left section) were considered “effective”. **B,** The schema showing the effect of STAG2 inactivation in DNA replication and PARP inhibitor in DNA damage repair (DDR) system. STAG2 inactivation induces stalled replication forks, leading to their collapse and double-strand DNA break (DDB). PARP inhibitor blocks DDR of single-strand DNA break and causes DDB. DDR for DDB is also hindered by PARP inhibitors due to PARP1-DNA trapping. HR, homologous recombination; NHEJ, non-homologous end joining. **C,** The dose-response curves of *STAG2/LMO2* T-ALL lines (MOLT-14 and PER-117) and STAG2 wild-type T-ALL lines (PEER and LOUCY) treated with PARP inhibitor, talazoparib in triplicate. EC_50_ was calculated after 48 hours of treatment. **D,** The synergistic effect of low-dose HDAC inhibitor, vorinostat (0.1 μM and 0.2 μM), with talazoparib was shown in the dose-response curves and in the highest single agent (HSA) synergy score in PER-117 (n=3). **E,** LC_50_ value of patient-derived xenografts (PDX) cells from γδ TCR positive *STAG2/LMO2* T-ALL tested by a panel of 26 drugs in duplicate. LC_50_ values for each tested drug in reference T-ALL cell lines were shown in black lines. LC_50_ was calculated after 96 hours of treatment. **F,** The dose-response curves of PDX cells with STAG2/LMO2 subtype treated by talazoparib and prednisolone for 96 hours in duplicate.

Recent studies have reported the efficacy of Poly(ADP-ribose) polymerase (PARP) inhibitors in STAG2-inactivated cancers, including glioblastoma, myelodysplastic syndromes (MDS), and acute myeloid leukemia (AML) due to stalled replication forks and increased double-strand DNA breaks^25–28^ (DDB, **Fig 5B**). The PARP inhibitor talazoparib was one of the most effective compounds in the *STAG2/LMO2*, but not *STAG2* wild-type T-ALL cell lines (**Fig 5C**). Moreover, synergism with a low dose HDAC inhibitor,^29,30^ vorinostat (0.1 and 0.2 µM), was observed in *STAG2/LMO2* lines, but not in *STAG2* wild-type lines (**Fig 5D** and **Appendix Fig A8F,G**).

Finally, we validated the drug efficacy on *STAG2/LMO2* subtype by using patient-derived xenografts (PDX) cells from γδ TCR positive T-ALL exposed to a panel of 26 drugs including conventional chemotherapeutics and pathway-directed agents^31^ (**Appendix Table A19**). This showed resistance to prednisolone that is consistent with slow treatment response in patients (**Fig 5E,F**). Effective drugs included PARP (talazoparib, olaparib), HDAC (vorinostat), BCL2 (venetoclax, navitoclax) and, BRD (JQ1) inhibitors, nelarabine and L-asparaginase, supporting the results of cell line screening (**Fig 5E,F**).

## DISCUSSION

Despite the recent improvement of outcomes in pediatric T-ALL,^1–3^ γδ T-ALL continues to pose a significant high-risk challenge. Due to the rarity of this disease and the lack of routine αβ/γδ TCR tests by flow cytometry at diagnosis in some clinical trials, comprehensive clinical and genomic analyses of γδ T-ALL have been lacking. To address this, we established a multi-national consortium that assembled a cohort of 200 pediatric γδ T-ALL patients across various study groups. These patients were compared with a group of 1,067 protocol-matched non-γδ T-ALL patients, shedding light on several distinct features of γδ T-ALL. One pivotal characteristic was the sluggish response to treatment and resistance to current therapeutic approaches. This resistance often necessitated intensified treatment regimens, including transplantation, and resulted in a high rate of mortality from both primary disease and treatment-related toxicity. Additionally, predominant BM relapses were observed, though CNS-related relapses are common in non-γδ T-ALL. Notably, these features were especially pronounced in patients under three years old or those with MRD ≥1% at EOI. These patient groups are considered extremely high-risk in the context of γδ T-ALL, necessitating alternative therapeutic strategies rather than treatment intensification. In contrast, other γδ T-ALL cases showed favorable outcomes and may continue to benefit from the current treatment strategy.

Our genomic analysis revealed that the *STAG2/LMO2* subtype was enriched in high-risk γδ T-ALL. The *STAG2/LMO2* subtype was predominantly associated with a younger age group, primarily affecting children under the age of three including infants. This subtype exhibits a distinct expression profile with dual-hit alterations of *LMO2* and *STAG2*. The young onset of *STAG2/LMO2* T-ALL, up-regulation of *HBE1* (the hemoglobin subunit expressed in the embryonic yolk sac), and the LIN28/let-7 pathway (whose expression in normal HSCs is limited to the fetal period, as well as γδ thymocytes^20,32^), may suggest a fetal hematopoietic origin. In addition, *Lin28b* expression was the highest in γδ T-cells among mouse thymocyte subsets at birth, and ectopic *Lin28b* expression in adult mouse HSCs promoted γδ T-cell development,^20,33^ supporting the enrichment of *STAG2/LMO2* ALL in γδ T-ALL resulting from the activation of LIN28/let-7 pathway in leukemogenesis.

Our analyses suggest that many of the distinct biologic features of *STAG2/LMO2* ALL may be attributed to inactivation of STAG2, considering the frequent activation of *LMO2* and *NOTCH1* across the landscape of T-ALL.^10,34–36^ STAG1 and STAG2 constitute a cohesin complex that is crucial in the maintenance of three-dimensional genome architecture^37^. Although partly redundant, these paralogues have distinct roles in chromatin accessibility and organization.^21,38–40^ STAG1 mediates demarcation of topologically associating domains (TAD) with CTCF, whereas STAG2 patterns H3K27ac and regulates cell-type specific transcription through short loops within TAD ^24,39–41^. The function of STAG2-cohesin is more prominent in generating loops between enhancers and promoters.^22–24,40^ Therefore, STAG2 inactivation alters enhancer-promoter looping, resulting in deregulation of transcription.^24,41,42^ Larger H3K27ac-based loops in *STAG2/LMO2* ALL and changes of H3K27ac peaks and expression at the regions of T-cell differentiation-related genes in our *STAG2* gene-edited models may suggest involvement of STAG2 inactivation in leukemogenesis by introducing differentiation arrest.

Inactivation of STAG2 may generate an “Achilles heel” in *STAG2/LMO2* ALL by increasing stalled and collapsed replication forks, leading to DDB.^27,28^ PARP inhibitors generate DDB by blocking DNA repair pathways and might accelerate the accumulation of DDB in *STAG2/LMO2* ALL, resulting in cell death, as seen in MDS/AML with alterations of *STAG2.*^25,28^ Talazoparib is a potent PARP inhibitor due to its strong PARP trapping,^43^ and was one of the most potent of 2,050 compounds in inhibiting *STAG2/LMO2* ALL cell proliferation, and its effect was specific to the *STAG2/LMO2* ALL. However, unlike *BRCA*-deficient cells, *STAG2/LMO2* ALL does not result in synthetic lethality by inhibition of single-strand DNA repair pathways through PARP inhibitors,^44,45^ because homologous recombination is not affected in *STAG2/LMO2* ALL, and thus the sensitivity of PARP inhibitors may depend on the strength of PARP trapping. Therefore, exploring combinatorial therapy is warranted to most effectively treat *STAG2/LMO2* ALL. HDAC inhibitors are attractive due to their observed efficacy, and their ability to inhibit double-strand DNA repair pathways via promoting PARP trapping through PARP1 accetylation.^29^

In conclusion, early onset γδ T-ALL observed in infants and very young children represents an exceptionally high-risk group of leukemia, prominently associated with *STAG2/LMO2* subtype. STAG2 inactivation perturbs chromatin organization and hematopoietic differentiation, whereas also rendering DNA repair pathways vulnerable. The insights gained from our findings hold the potential to inform more precise refined risk stratification at diagnosis and pave the way for innovative therapeutic strategies that are needed to cure this form of leukemia.

## Author information

### Author contributions

A.Y., A.A., A.S.M., A.M., B.B., C.K., C.L., C.H.P., D.T., E.M., E.V., F.L., G.E., H.E., H.M., H.D., J.R., J.S., J.C., J.M., J.Takita, K.P., K.S., M.E., M.Schrappe, M.Z., M.T., M.M., M.R., M.Kicinski, M.Kato, O.S., P.M., R.M., R.K., R.P., S.E., S.Köhrer, S.L., S.Shen, S.J., S.M.K., S.Skoczeń, T.M., T.V., T.I., V.C., Y.T., K.R., P.T., D.T.T. provided patient samples and data. S.Kimura, H.I., K.Caldwell, Y.L., K.H., Z.Y., C.Cheng analyzed clinical data. S.Kimura, D.T.T., P.P., L.M., C.Chen, Z.C., C.Q., J.Trka, M.Svaton, Q.G., Z.G. generated and analyzed genomic data. S.Kimura, A.B., E.D.A., Y.C., I.I., B.F., C.S.P., S.M., M.S.P., S.M.P., N.R., K.Crews, J.Y. conducted experiments. S.Kimura, H.I., C.G.M. designed the study, oversaw experiments, and wrote the manuscript.

## Data Availability

Primary sample whole-genome and transcriptome data have been deposited in the European Genome-phenome Archive (EGA) with accession number EGAS50000000018, in the database of genotypes and phenotypes (dbGaP; http://www.ncbi.nlm.nih.gov/gap) under accession number phs002276.v2.p1 (phs000218, phs000464), in the Kids First data portal (https://portal.kidsfirstdrc.org/dashboard), and in the National Bioscience Database Center (NBDC) database with accession number JGAS000090. HiChIP and ChIP-seq data have been deposited at GSE243776. Reference ChIP-seq and HiChIP data used in this study have been deposited at EGAS50000000016 and GSE165209.

https://www.ncbi.nlm.nih.gov/geo/query/acc.cgi?acc=GSE243776

## ACKNOWLEDGMENTS

Fig 4B, Fig 5B, Appendix Fig 3B, and Supplemental Figure 1a were created with BioRender.com.

## Competing interests

D.T.T. received research funding from BEAM Therapeutics, NeoImmune Tech and serves on advisory boards for BEAM Therapeutics, Janssen, Servier, Sobi, and Jazz. D.T.T. has multiple patents pending on CAR-T. I.I. received honoraria from Amgen and Mission Bio. M.Kicinski received research funding from MSD, BMS, Pierre Fabre, JnJ, and Immunocore. C.G.M. received research funding from Loxo Oncology, Pfizer, AbbVie; honoraria from Amgen and Illumina, and holds stock in Amgen. There are no conflicts of interest with the work presented in this manuscript.

## Notes

### Author Declarations

All clinical trials were approved by institutional review boards or ethics committees. This study was ethically approved by the Institutional Review Board of St. Jude Children's Research Hospital.

## REFERENCES

1. Winter SS, Dunsmore KP, Devidas M, et al: Improved Survival for Children and Young Adults With T-Lineage Acute Lymphoblastic Leukemia: Results From the Children’s Oncology Group AALL0434 Methotrexate Randomization. J Clin Oncol 36:2926–2934, 2018

2. Dunsmore KP, Winter SS, Devidas M, et al: Children’s Oncology Group AALL0434: A Phase III Randomized Clinical Trial Testing Nelarabine in Newly Diagnosed T-Cell Acute Lymphoblastic Leukemia. J Clin Oncol 38:3282–3293, 2020

3. Sato A, Hatta Y, Imai C, et al: Nelarabine, intensive L-asparaginase, and protracted intrathecal therapy for newly diagnosed T-cell acute lymphoblastic leukaemia in children and young adults (ALL-T11): a nationwide, multicenter, phase 2 trial including randomisation in the very high-risk group. Lancet Haematol 10:e419–e432, 2023

4. Schrappe M, Valsecchi MG, Bartram CR, et al: Late MRD response determines relapse risk overall and in subsets of childhood T-cell ALL: results of the AIEOP-BFM-ALL 2000 study. Blood 118:2077–84, 2011

5. Pui CH, Pei D, Cheng C, et al: Treatment response and outcome of children with T-cell acute lymphoblastic leukemia expressing the gamma-delta T-cell receptor. Oncoimmunology 8:1599637, 2019

6. Matos DM, Rizzatti EG, Fernandes M, et al: Gammadelta and alphabeta T-cell acute lymphoblastic leukemia: comparison of their clinical and immunophenotypic features. Haematologica 90:264–6, 2005

7. Wieduwilt MJ: Ph+ ALL in 2022: is there an optimal approach? Hematology Am Soc Hematol Educ Program 2022:206–212, 2022

8. Inaba H, Mullighan CG: Pediatric acute lymphoblastic leukemia. Haematologica 105:2524–2539, 2020

9. Tanasi I, Ba I, Sirvent N, et al: Efficacy of tyrosine kinase inhibitors in Ph-like acute lymphoblastic leukemia harboring ABL-class rearrangements. Blood 134:1351–1355, 2019

10. Liu Y, Easton J, Shao Y, et al: The genomic landscape of pediatric and young adult T-lineage acute lymphoblastic leukemia. Nat Genet 49:1211–1218, 2017

11. Seki M, Kimura S, Isobe T, et al: Recurrent SPI1 (PU.1) fusions in high-risk pediatric T cell acute lymphoblastic leukemia. Nat Genet 49:1274–1281, 2017

12. Montefiori LE, Bendig S, Gu Z, et al: Enhancer Hijacking Drives Oncogenic BCL11B Expression in Lineage-Ambiguous Stem Cell Leukemia. Cancer Discov 11:2846–2867, 2021

13. Kimura S, Montefiori L, Iacobucci I, et al: Enhancer retargeting of CDX2 and UBTF::ATXN7L3 define a subtype of high-risk B-progenitor acute lymphoblastic leukemia. Blood 139:3519–3531, 2022

14. Pieters R, de Groot-Kruseman H, Van der Velden V, et al: Successful Therapy Reduction and Intensification for Childhood Acute Lymphoblastic Leukemia Based on Minimal Residual Disease Monitoring: Study ALL10 From the Dutch Childhood Oncology Group. J Clin Oncol 34:2591–601, 2016

15. Olivier-Gougenheim L, Arfeuille C, Suciu S, et al: Pediatric randomized trial EORTC CLG 58951: Outcome for adolescent population with acute lymphoblastic leukemia. Hematol Oncol 38:763–772, 2020

16. Pui CH, Campana D, Pei D, et al: Treating childhood acute lymphoblastic leukemia without cranial irradiation. N Engl J Med 360:2730–41, 2009

17. Jeha S, Pei D, Choi J, et al: Improved CNS Control of Childhood Acute Lymphoblastic Leukemia Without Cranial Irradiation: St Jude Total Therapy Study 16. J Clin Oncol 37:3377–3391, 2019

18. Buchmann S, Schrappe M, Baruchel A, et al: Remission, treatment failure, and relapse in pediatric ALL: an international consensus of the Ponte-di-Legno Consortium. Blood 139:1785–1793, 2022

19. Pölönen P, Elsayed A, Di Giacomo D, et al: Comprehensive Genome Characterization of Childhood T-ALL Links Oncogene Activation Mechanism and Subtypes to Prognosis. Blood 140:1727–1729, 2022

20. Yuan J, Nguyen CK, Liu X, et al: Lin28b reprograms adult bone marrow hematopoietic progenitors to mediate fetal-like lymphopoiesis. Science 335:1195–200, 2012

21. Cuadrado A, Losada A: Specialized functions of cohesins STAG1 and STAG2 in 3D genome architecture. Curr Opin Genet Dev 61:9–16, 2020

22. Adane B, Alexe G, Seong BKA, et al: STAG2 loss rewires oncogenic and developmental programs to promote metastasis in Ewing sarcoma. Cancer Cell 39:827–844 e10, 2021

23. Surdez D, Zaidi S, Grossetete S, et al: STAG2 mutations alter CTCF-anchored loop extrusion, reduce cis-regulatory interactions and EWSR1-FLI1 activity in Ewing sarcoma. Cancer Cell 39:810–826 e9, 2021

24. Viny AD, Bowman RL, Liu Y, et al: Cohesin Members Stag1 and Stag2 Display Distinct Roles in Chromatin Accessibility and Topological Control of HSC Self-Renewal and Differentiation. Cell Stem Cell 25:682–696 e8, 2019

25. Tothova Z, Valton AL, Gorelov RA, et al: Cohesin mutations alter DNA damage repair and chromatin structure and create therapeutic vulnerabilities in MDS/AML. JCI Insight 6, 2021

26. Bailey ML, O’Neil NJ, van Pel DM, et al: Glioblastoma cells containing mutations in the cohesin component STAG2 are sensitive to PARP inhibition. Mol Cancer Ther 13:724–32, 2014

27. Padella A, Ghelli Luserna Di Rora A, Marconi G, et al: Targeting PARP proteins in acute leukemia: DNA damage response inhibition and therapeutic strategies. J Hematol Oncol 15:10, 2022

28. Mondal G, Stevers M, Goode B, et al: A requirement for STAG2 in replication fork progression creates a targetable synthetic lethality in cohesin-mutant cancers. Nat Commun 10:1686, 2019

29. Robert C, Nagaria PK, Pawar N, et al: Histone deacetylase inhibitors decrease NHEJ both by acetylation of repair factors and trapping of PARP1 at DNA double-strand breaks in chromatin. Leuk Res 45:14–23, 2016

30. Kruglov O, Wu X, Hwang ST, et al: The synergistic proapoptotic effect of PARP-1 and HDAC inhibition in cutaneous T-cell lymphoma is mediated via Blimp-1. Blood Adv 4:4788–4797, 2020

31. Rowland L, Smart B, Brown A, et al: Ex vivo Drug Sensitivity Imaging-based Platform for Primary Acute Lymphoblastic Leukemia Cells. Bio Protoc 13:e4731, 2023

32. Tieppo P, Papadopoulou M, Gatti D, et al: The human fetal thymus generates invariant effector gammadelta T cells. J Exp Med 217, 2020

33. Dong M, Mallet Gauthier E, Fournier M, et al: Developing the right tools for the job: Lin28 regulation of early life T-cell development and function. FEBS J 289:4416–4429, 2022

34. Abdulla HD, Alserihi R, Flensburg C, et al: Overexpression of Lmo2 initiates T-lymphoblastic leukemia via impaired thymocyte competition. J Exp Med 220, 2023

35. Sanchez-Martin M, Ferrando A: The NOTCH1-MYC highway toward T-cell acute lymphoblastic leukemia. Blood 129:1124–1133, 2017

36. McCormack MP, Young LF, Vasudevan S, et al: The Lmo2 oncogene initiates leukemia in mice by inducing thymocyte self-renewal. Science 327:879–83, 2010

37. Davidson IF, Bauer B, Goetz D, et al: DNA loop extrusion by human cohesin. Science 366:1338–1345, 2019

38. Waldman T: Emerging themes in cohesin cancer biology. Nat Rev Cancer 20:504–515, 2020

39. Casa V, Moronta Gines M, Gade Gusmao E, et al: Redundant and specific roles of cohesin STAG subunits in chromatin looping and transcriptional control. Genome Res 30:515–527, 2020

40. Kojic A, Cuadrado A, De Koninck M, et al: Distinct roles of cohesin-SA1 and cohesin-SA2 in 3D chromosome organization. Nat Struct Mol Biol 25:496–504, 2018

41. Smith JS, Lappin KM, Craig SG, et al: Chronic loss of STAG2 leads to altered chromatin structure contributing to de-regulated transcription in AML. J Transl Med 18:339, 2020

42. Chu Z, Gu L, Hu Y, et al: STAG2 regulates interferon signaling in melanoma via enhancer loop reprogramming. Nat Commun 13:1859, 2022

43. Muvarak NE, Chowdhury K, Xia L, et al: Enhancing the Cytotoxic Effects of PARP Inhibitors with DNA Demethylating Agents - A Potential Therapy for Cancer. Cancer Cell 30:637–650, 2016

44. Farmer H, McCabe N, Lord CJ, et al: Targeting the DNA repair defect in BRCA mutant cells as a therapeutic strategy. Nature 434:917–21, 2005

45. Bryant HE, Schultz N, Thomas HD, et al: Specific killing of BRCA2-deficient tumours with inhibitors of poly(ADP-ribose) polymerase. Nature 434:913–7, 2005

