## Appendix Fig for "Biologic and clinical features of childhood gamma delta T-ALL: identification of STAG2/LMO2 γδ T-ALL as an extremely high risk leukemia in the very young"

### **Appendix Figures**

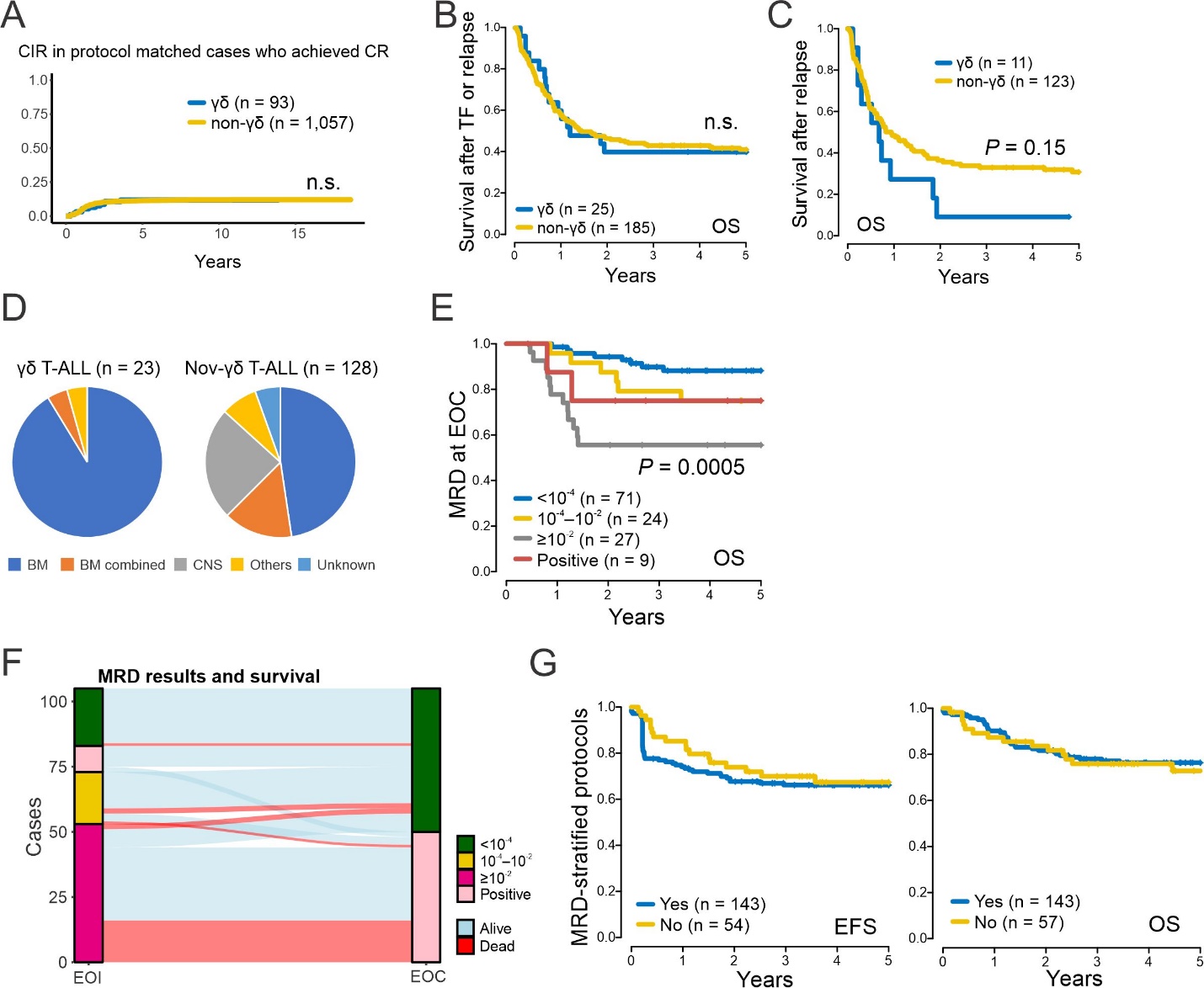

#### **Appendix Figure A1. The clinical features of γδ T-ALL.** **A,** The cumulative incidence of relapse CIR) in patients with γδ T-ALL (n = 93) compared to those of non-γδ T-ALL (n = 1,057) is shown. No difference in CIR was observed among cases in protocol-matched cohorts. n.s., not significant. **B,** Overall survival (OS) after the first events comparing γδ T-ALL and non-γδ T-ALL with treatment failure (TF) or relapse. **C,** OS after relapses comparing γδ T-ALL and non-γδ T-ALL. **D,** Relapse sites in γδ T-ALL (n = 23) and non-γδ T-ALL (n = 128). Bone marrow (BM) was the most frequent relapse site for γδ T-ALL, while central nervous system (CNS) related relapse (including BM combined) accounted for one-third of non-γδ T-ALL relapse sites. **E,** γδ T-ALL patients with minimal residual disease (MRD) >1% at the end of consolidation (EOC) exhibited significantly worse OS. **F,** Changes of MRD status at the end of induction (EOI) and EOC with outcomes in γδ T-ALL were shown. Most of MRD >1% cases at EOI remained MRD positive (>0.01%) at EOC and almost half died. **G,** Event-free survival (EFS) and OS of γδ T-ALL comparing cases that received MRD-stratified protocols. MRD-stratified protocols did not improve outcomes of γδ T-ALL.

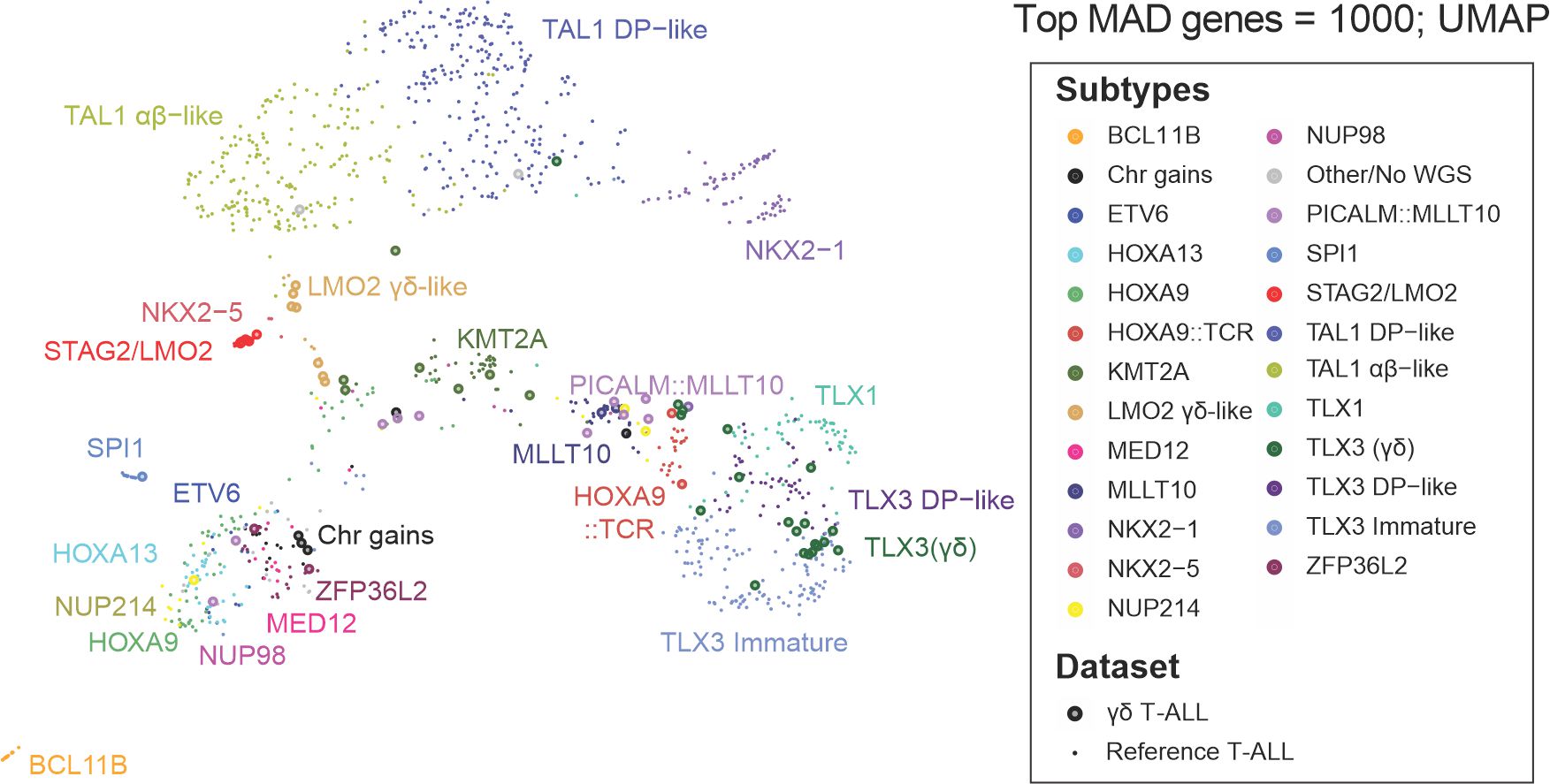

#### **Appendix Figure A2. The gene expression profiles of γδ T-ALL.** UMAP plot of gene expression analysis for 68 cases of γδ T-ALL (large donut shape circles) layered on reference T-ALL cohort (n = 1,076, small circles), of which genomic subtypes were known, by using the top 1,000 most variable genes. γδ T-ALL cases were classified not by T-cell receptor (TCR) lineages (αβ, γδ, and no TCR) but by leukemia-initiating events.

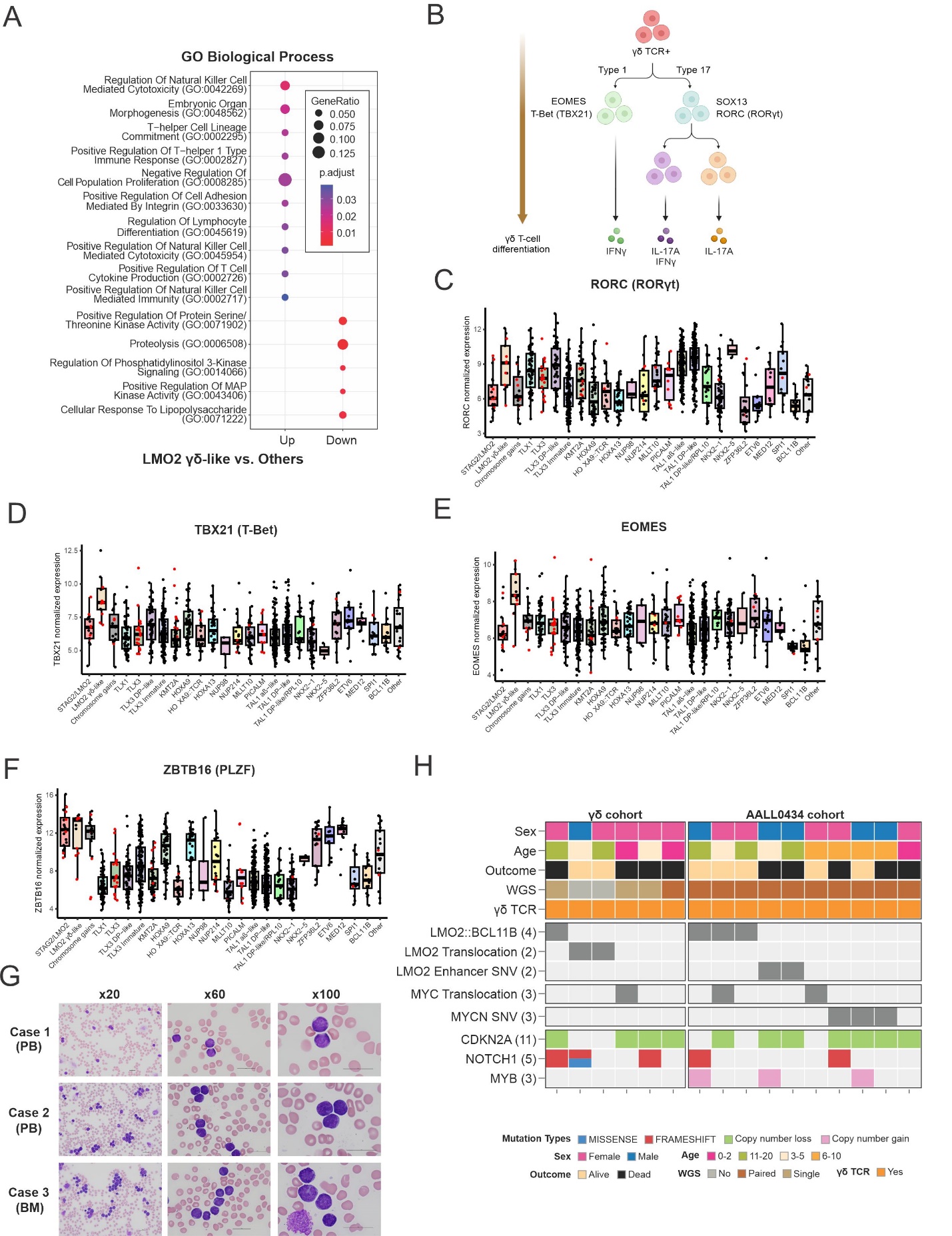

#### **Appendix Figure A3. The genomic feature of *LMO2* γδ-like** **T-ALL.** **A,** Pathway analysis (GO Biological Process) using up-regulated genes (n=70, adjusted *P*<0.01 and fold change >2) in *LMO2* γδ-like compared to other T-ALL cases. **B,** The schema of normal γδ T-cell differentiation after the bifurcation at CD4/CD8 double negative stage^1,2^. Thymocytes expressing γδ T-cell receptor (TCR) bifurcate into IFNγ-producing Type 1 γδ T-cells and IL17-producing Type 17 γδ T-cells with expression of characteristic genes. The normalized expression level of **(C)** *RORC*, **(D)** *TBX21*, **(E)** *EOMES*, and **(F)** *ZBTB16* comparing each genomic subtype of T-ALL. *LMO2* γδ-like subtype is shown at the second from the left in light brown. γδ T-ALL cases are shown in red color. **G,** The peripheral blood (PB) or bone marrow (BM) smear of three available *LMO2* γδ-like cases at diagnosis. In all cases, leukemic blast cells included more mature features with small and/or chromatin-condensation. The scale bar indicates 20 µm. **H,** The heatmap showing the mutational landscape and clinical parameters of the 16 cases of *LMO2* γδ-like T-ALL in γδ T-ALL cohort (n = 6) and reference cohort (n = 10). *LMO2*, *MYC*, and *MYCN* alterations were found in 13 out of 16 cases with almost mutually exclusive patterns.

1.	Serre K, Silva-Santos B: Molecular Mechanisms of Differentiation of Murine Pro-Inflammatory gammadelta T Cell Subsets. Front Immunol 4:431, 2013

2.	Munoz-Ruiz M, Sumaria N, Pennington DJ, et al: Thymic Determinants of gammadelta T Cell Differentiation. Trends Immunol 38:336-344, 2017

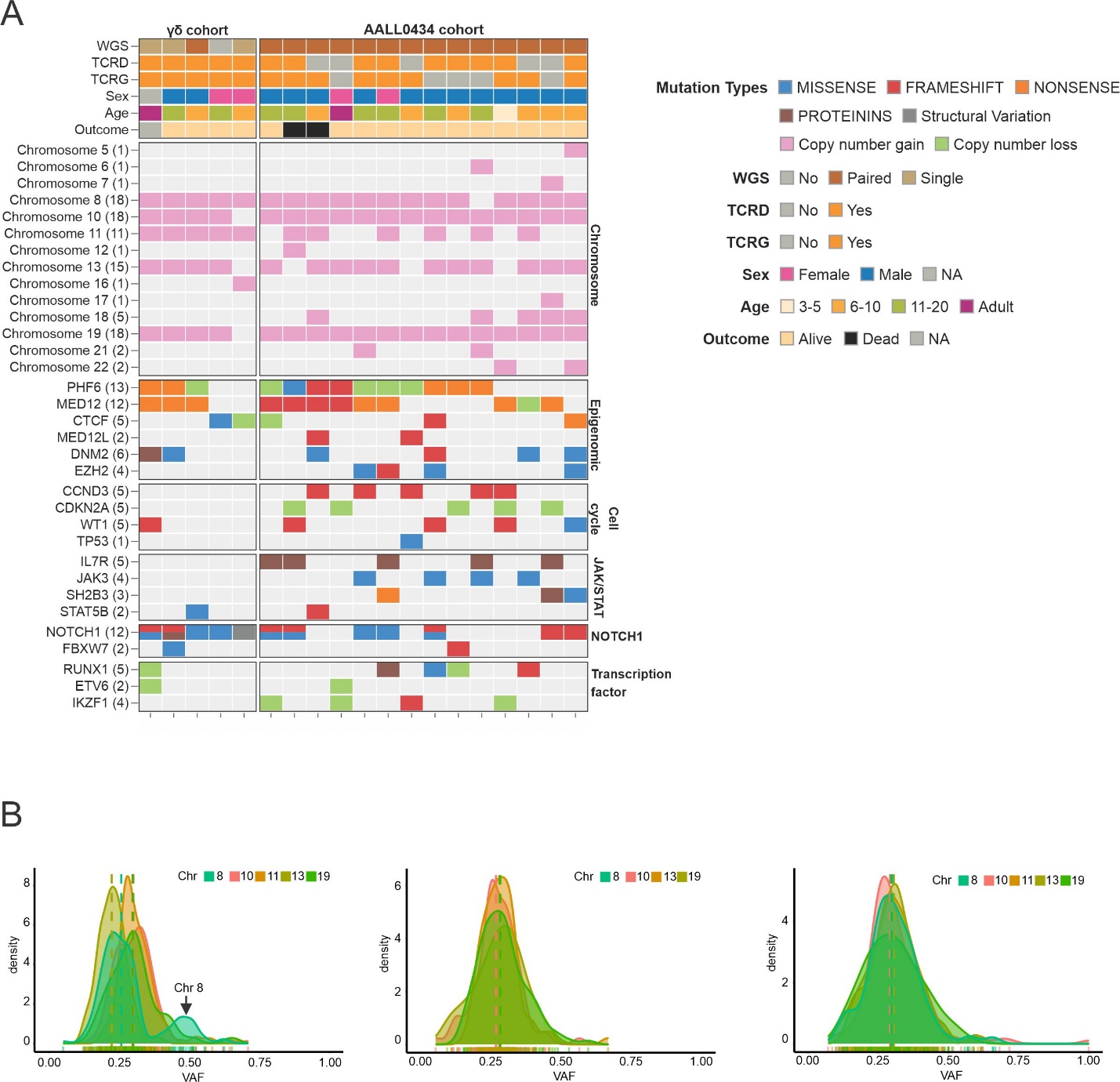

#### **Appendix Figure A4. The genomic feature of Chromosome gains subtype.** **A,** The heatmap showing the patterns of chromosome gain, mutational landscape, and clinical parameters of the 19 cases of hyperdiploid T-ALL, Chromosome gains subtype, in γδ T-ALL cohort (n = 5) and reference cohort (n = 14). Recurrent gains of +8, +10, +11, +13q, +19 chromosomes were detected in most cases. All cases had one or more of *PHF6*, *MED12*, and *CTCF* alterations. **B,** Density plots showing whether copy gains are likely to have occurred simultaneously or sequentially by using variant allele frequency (VAF) of somatic single nucleotide variants from whole genome sequence data. Only one case exhibited an asynchronous gain of chromosome 8 (arrow); the existence of mutations with VAFs around 0.5 indicates late copy gains because these mutations were acquired before copy gains. In all the other 14 cases, the timing of the acquisition of aneuploidies was synchronous (representative 2 cases are shown).

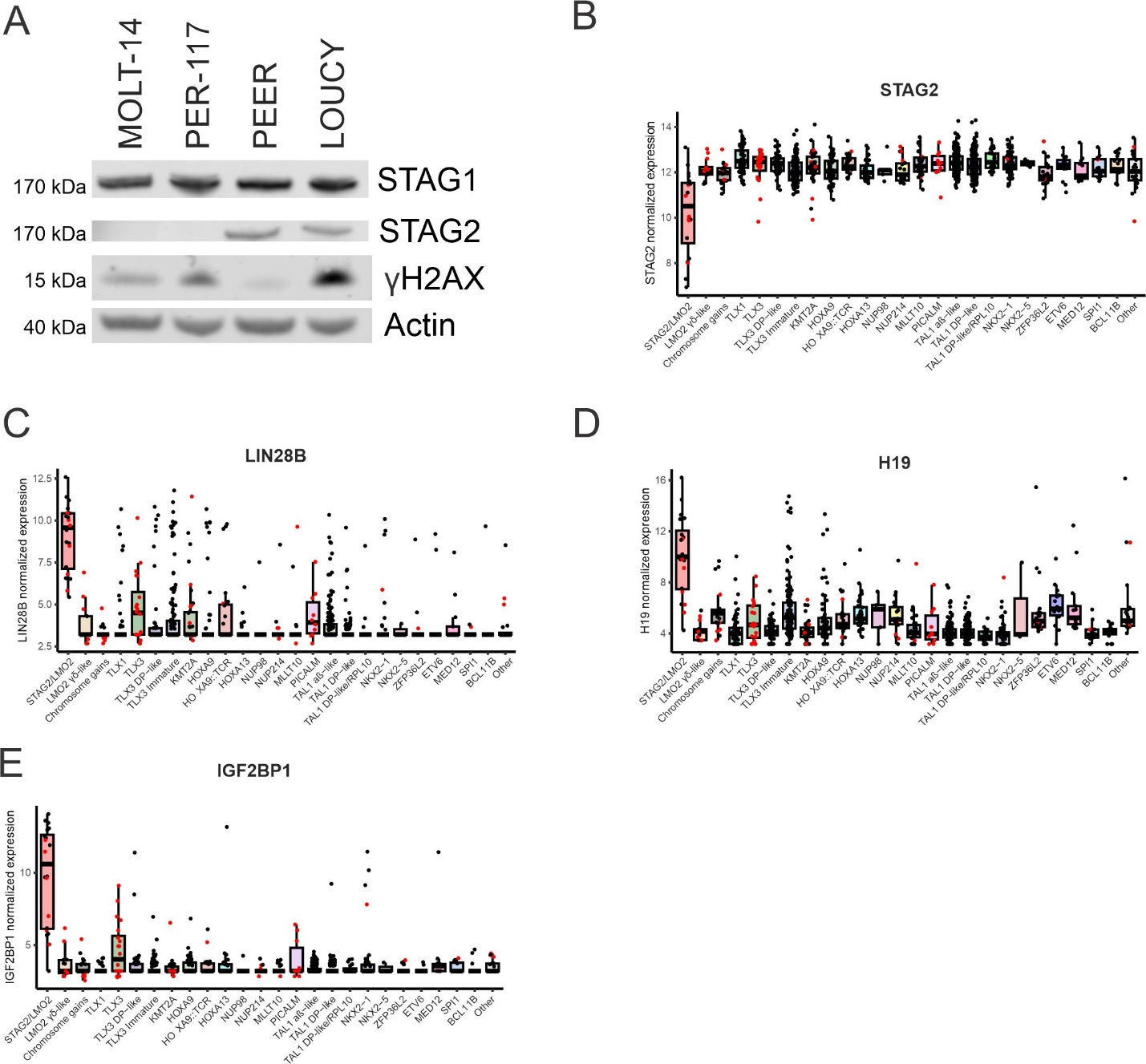

#### **Appendix Figure A5. The feature gene expression of *STAG2/LMO2* subtype T-ALL.** **A,** Immunoblotting showing STAG1, STAG2, and Phospho-Histone H2A.X (Ser139) (γH2AX) proteins in STAG2/LMO2 subtype cell lines (MOLT-14 and PER-117), non STAG2/LMO2 cell lines, PEER (γδ T-ALL) and LOUCY (early T-cell precursor ALL). STAG2 was not expressed in both of *STAG2/LMO2* subtype cell lines. The normalized expression level of **(B)** *STAG2*, **(C)** *LIN28B*, **(D)** *H19*, and **(E)** *IGF2BP1* comparing each genomic subtype of T-ALL is shown. *STAG2/LMO2* subtype is shown at the left in light red. γδ T-ALL cases are shown in red color.

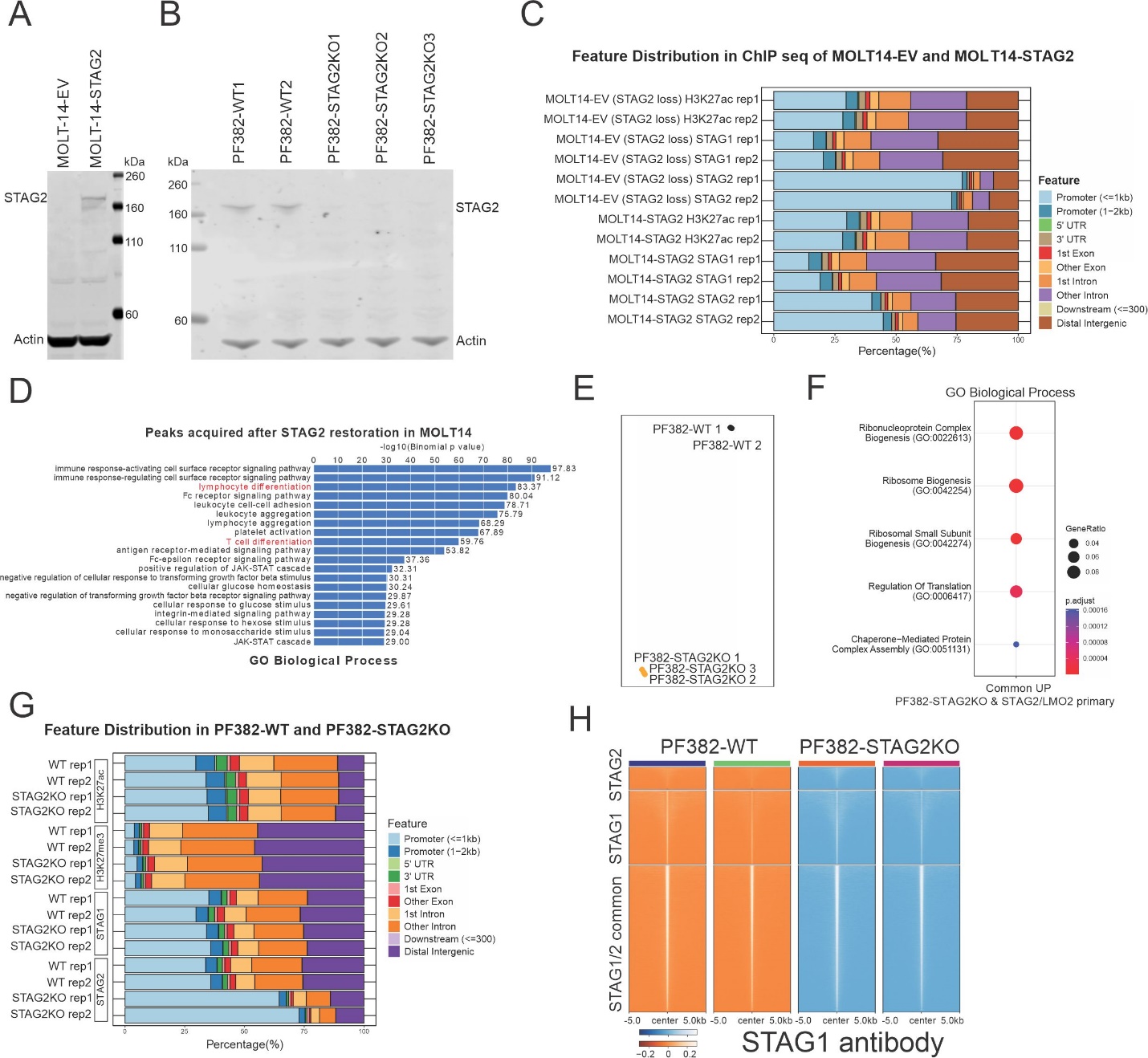

#### **Appendix Figure A6. Models for examining the effects of STAG2 inactivation.** Immunoblotting showing STAG2 proteins in **(A)** STAG2 restoration model (MOLT14-EV and MOLT14-STAG2) and **(B)** STAG2 knockout (KO) model (PF382-WT and PF382-STAG2KO). **C,** Distribution of H3K27ac ChIP-seq peaks in replicates (rep) 1 and 2 of MOLT14-EV and MOLT14-STAG2 across genomic regions. **D,** Pathway analysis (GO Biological Process) using H3K27ac peaks that appeared after STAG2 restoration from MOLT14 having *LMO2::STAG2* (peaks possibly lost by STAG2 inactivation) showed enrichment of T-cell differentiation-related pathways. **E,** Gene expression profiling of gene-edited PF382 cells (WT and STAG2KO) represented in UMAP plot. STAG2 KO induced the distinct expression profiles. **F,** Pathway analysis (GO Biological Process) using common up-regulated genes (adjusted P <0.01) in PF382-STAG2KO v. PF382-WT and primary samples with *STAG2/LMO2* subtypes v. other T-ALL subtype samples. Ribosome biogenesis pathways were enriched. **G,** Distribution of H3K27ac ChIP-seq peaks in replicates (rep) 1 and 2 of PF382-WT and PF382-STAG2KO across genomic regions. **H,** ChIP-seq density heatmap of STAG1 in PF382-WT and PF382-STAG2KO lines around STAG1/STAG2 common, STAG1, and STAG2 binding regions detected from PF382-WT.

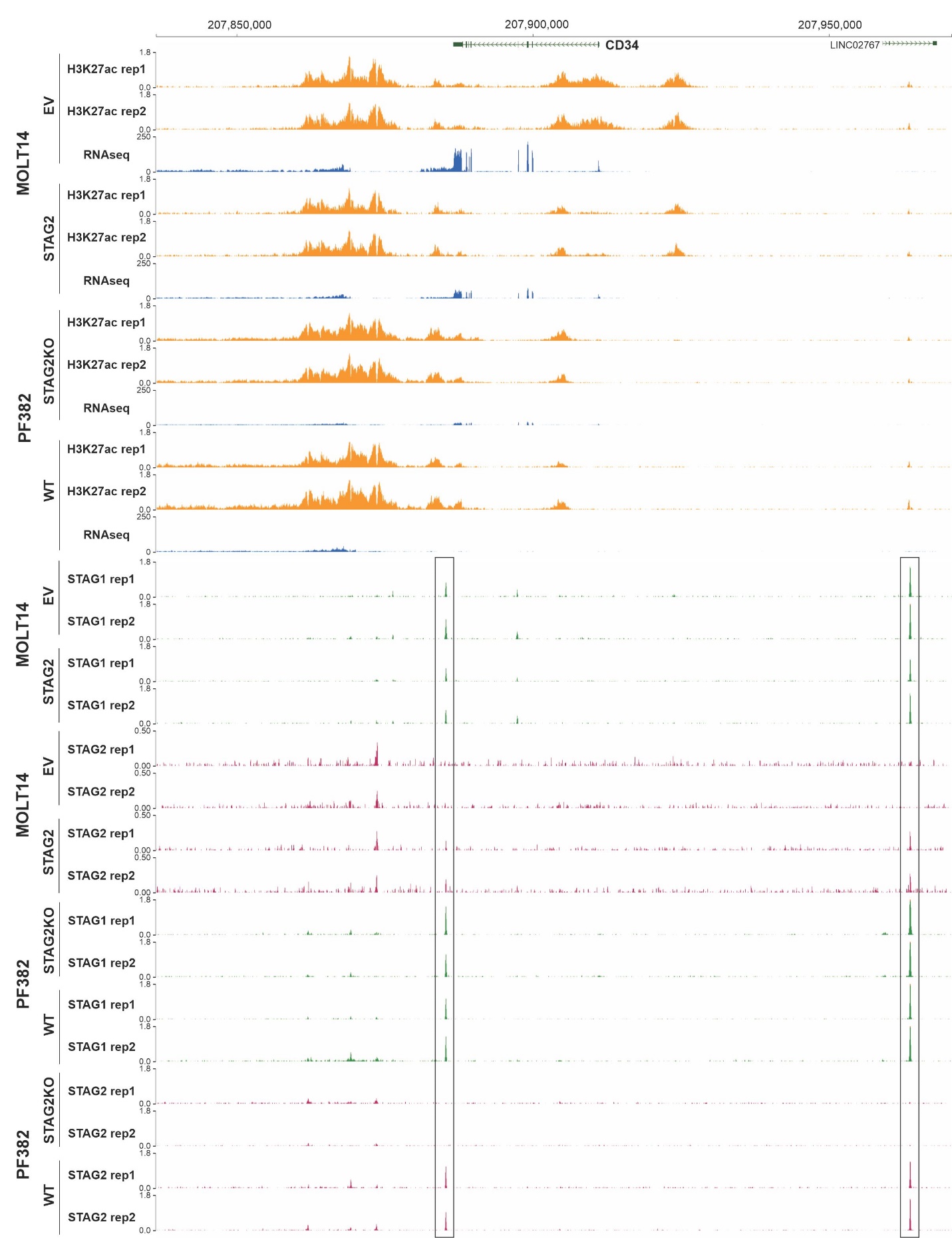

#### **Appendix Figure A7. ChIP-seq coverage at the CD34 locus showing the effects of STAG2 inactivation.** H3K27ac (orange), STAG1 (green), and STAG2 (red) binding and RNAseq (blue) coverage at the *CD34* locus in MOLT14-EV, MOLT14-STAG2, PF382-WT, and PF382-STAG2KO cells in duplicates. The black squares indicate deregulated STAG2 binding sites.

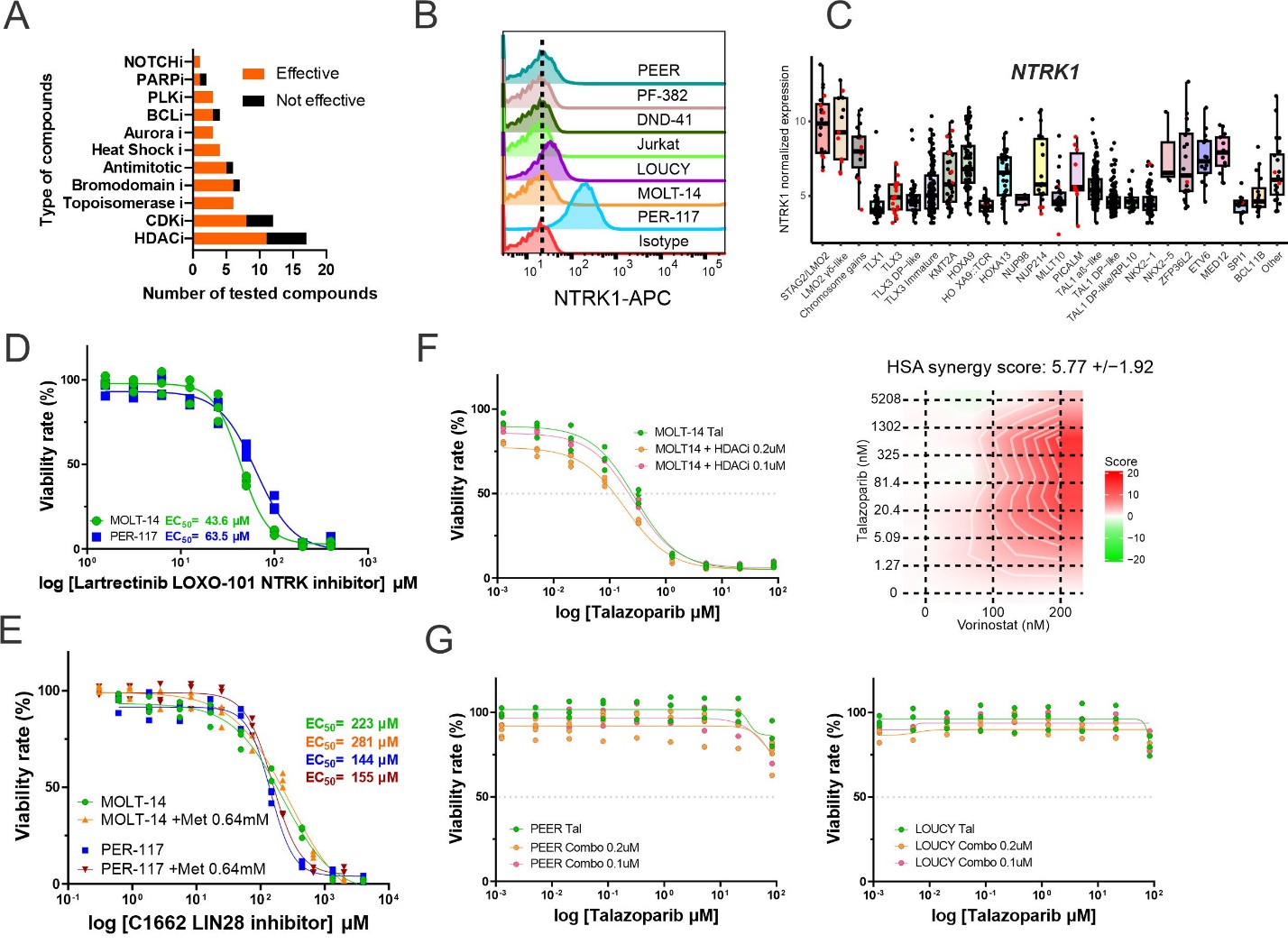

#### **Appendix Figure A8. Exploration of targets in *STAG2/LMO2* T-ALL.** **A,** The number of effective and non-effective compounds included in each pathway from the result of the dose-response analysis for 138 compounds. **B,** NTRK1 expression in each T-ALL cell line assessed by flow cytometry. PER-117 (*STAG2/LMO2*) expressed NTRK1. **C,** The normalized expression level of *NTRK1* comparing each genomic subtype of T-ALL is shown. *STAG2/LMO2* subtype is shown at the left in light red. γδ T-ALL cases are shown in red color. **D,** The dose-response curves of *STAG2/LMO2* lines, MOLT-14 (NTRK1 negative) and PER-117 (NTRK1 positive) treated with NTRK1 inhibitor, lartrectinib (LOXO-101). EC_50_ was calculated after 48 hours of treatment. Lartrectinib was not effective in *STAG2/LMO2* lines regardless of NTRK1 expression. **E,** The dose-response curves of *STAG2/LMO2* lines treated with LIN28 inhibitor (C1662) with/without metformin (Met). EC_50_ was calculated after 48 hours of treatment. C1662 was not effective in *STAG2/LMO2* lines and no synergistic effect with Met was shown. **F,** The additive effect of low-dose HDAC inhibitor, vorinostat (0.1 μM and 0.2 μM), with talazoparib was shown in the dose-response curves and in the highest single agent (HSA) synergy score in MOLT-14 (n=3). **G,** The dose-response curves of *STAG2* wild-type T-ALL lines treated with talazoparib (Tal) with HDAC inhibitor vorinostat. Treatment (48 hours) of Tal and combination were not effective on these lines.
