## Supplementary material for "Biologic and clinical features of childhood gamma delta T-ALL: identification of STAG2/LMO2 γδ T-ALL as an extremely high risk leukemia in the very young": Data Supplement

### **SUPPLEMENTARY METHODS**

#### **Primary materials**

DNA, RNA, and primary materials of γδ T-ALL from the clinical study cohort were collected if available (N=61), with some extra material from patients with incomplete clinical data (N=5) and adult cases in the ECOG-ACRIN Cancer Research Group (ECOG-ACRIN, N=10). DNA and RNA were isolated from peripheral blood or BM samples according to standard protocols (AllPrep DNA/RNA Mini Kit, QIAGEN, # 80204 or Quick-DNA/RNA Microprep Plus Kit, ZYMO RESEARCH, #D7005). Cell sorting was performed for the materials with a low blast content (below 70%) by using the gate on T-ALL blast cells (CD45^dim^CD7^+^; CD45-FITC (2D1), BD, #340664, CD7-APC, BD, #MHCD0705, CD19-PE (4G7), BD, #349209; Data Supplement, **Supplementary Fig S2)**.

#### **Whole Genome Sequencing (WGS)**

In total WGS was performed for 47 γδ T-ALL samples with matched non-tumor samples for 19, and tumor only samples for 28. WGS data were processed according to our previous studies^1,2^. Briefly, Illumina-based sequencing reads were aligned to GRCh37 human genome reference by Burrows-Wheeler Alignment^3^ (BWA, version 0.7.17). For the mutation calls, we used the combination of (1) GenomonFisher (filtered out all calls < 1.15, version 0.2.1), GenomonMutationFilter (version 0.2.9), and EBFilter (filtered out all calls <4.5, version 0.2.1; <https://genomonproject.github.io/GenomonPagesR/>), and (2) GATK (version 4.1.2). For tumor-only samples, filtration was performed by using T-ALL-mutant-gene set which includes genes reported to have alterations in our previous T-ALL WES cohort^4^ (**Appendix Table A20**). The detection of structural variants (SVs) was performed by using the combination of four SV callers including GenomonSV (version 0.8.0), Delly^5^ (version 0.7.7), Lumpy (version 0.2.13), and Manta^6^ (version 1.6.0). All variant calls were further manually reviewed for read depth and to remove artifacts using the Integrated Genome Viewer^7^. CNVkit (version 0.9.1), conserting^8^, and cn.mops^9^ (version 1.30.0) were used for analyzing copy number alteration. Timing of copy number alterations in the cases with Chromosome gains were analyzed by using somatic single nucleotide variants (SNVs) on three-copy chromosomes from tumor-germline paired WGS according to our previous study^10^. Synchronous copy gains were defined when all three-copy chromosomes had a similar ratio of one of three (VAF=0.33) versus two of three (VAF=0.67) SNVs. Asynchronous copy gains were defined when the ratio was different for different chromosomes.

#### **Whole Transcriptome Sequencing (RNAseq)**

RNAseq data were processed according to our previous studies^11^. For some cases, RNAseq data were previously analyzed and publicly available^4,12^. Briefly, RNAseq data were aligned to the GRCh37 human genome reference by STAR^13^ (version 2.4.2a) using the suggested 2-pass mapping pipeline. For the detection of fusion genes, the combination of FusionCatcher (version 1.10; <https://github.com/ndaniel/fusioncatcher>) and STAR-Fusion (version 1.5.0) was used. To quantitate gene expression, HTSeq^14^ (version 0.13.5) was used to calculate read counts for each transcript with the following batch correction by ComBat function in the sva package^15^ as we previously described. The DESeq2 package^16^ was used for gene expression normalization and differentially expressed gene analysis. ClusterProfiler R package^17^ was used for the pathway analysis. For a 2-dimensional Uniform Manifold Approximation and Projection (UMAP) plot, we used top 1,000 most variable genes based on median absolute deviation or feature gene sets that can effectively describe T-ALL subtype from our previous study (**Appendix Table A8**). For cases without DNA, GATK (version 4.2.5.0) was used to detect gene alterations and filtered genes by using T-ALL-frequently-mutant-gene set (**Appendix Table A20**). For the analysis of copy number alteration, RNAseqCNV^18^ (<https://github.com/honzee/RNAseqCNV>) was used to estimate whole chromosome copy number alterations.

#### **HiChIP and ChIP-seq**

Cells were crosslinked by formaldehyde (Fisher Scientific, #F79-500) for 10 mins (2% for HiChIP and 1% for ChIP-seq) and quenched with 10X glycine (Cell Signaling, #7005) for 5 mins. HiChIP was performed by using the Arima-HiC+ kit (Arima, #A410231). Covaris was used for chromatin shearing and H3K27ac (Active Motif, #91193) and CTCF (Active Motif, #91285) antibodies were used for immunoprecipitation. HiChIP^19^ data including significant loops and loop size were processed and analyzed using MAPS software^20^ (version 2.0) as described in our previous study^1^. ChIP-seq was performed using the SimpleChIP Plus Sonication Chromatin IP kit (Cell Signaling, #56383) and the following antibodies were used for immunoprecipitation; H3K27ac (Active Motif, #91193), STAG1 (abcam, #ab4457), and STAG2 (abcam, #ab4463). ChIP-seq data were processed according to our previous study^21^. Briefly, sequencing reads were mapped to GRCh38 human genome reference by BWA^3^. MACS2^22^ (version 2.1.1) was used for peak calls. DiffBind R package was used for detecting differential binding peaks. Enriched gene sets were examined with GREAT^23^ (version 4.04).

#### **TCR repertoire analysis**

The fastq files of RNAseq and WGS data were aligned to reference V, D, J, and C genes of TCR and assembled clonotypes through MiXCR^24^ (version 3.0.13). The TCR reference used in the present study was included in the MiXCR software as default setting: TRA/TRD, NG_001332.2; TRB, NG_001333.2; TRG, NG_001336.2. TCR rearrangements calls were filtered by excluding (i) cloneCount < 5 and (ii) cloneFraction < 0.1 after the calculation of TCR-specific frequency. Amplicon TRD/G sequencing was used for the validation of the TCR repertoire for 20 representative cases. TRG and TRD sequencing libraries were prepared separately according to the protocols developed by the EuroClonality-NGS Working Group^25^, each from 70–100 ng of genomic DNA (corresponding to approximately 10,000-15,000 nucleated cells). Final libraries were sequenced on the MiSeq instrument with 2 x 250 v2 Reagent Kit (Illumina, San Diego, CA) and the data were analyzed with the ARResT/Interrogate platform^26^.

#### **Cell Culture**

The MOLT-14, PF-382, LOUCY, PEER, and 293T cells were purchased from the American Type Culture Collection (ATCC, catalog nos. CRL-3216 and CRL-2629) or Leibniz Institute DSMZ (catalog no. ACC 437, ACC38, and ACC6). PER-117 cells were kindly provided by Dr Rishi S Kotecha. All cell lines were authenticated by DNA profiling (short tandem repeat). 293T cells were cultured in Dulbecco’s modified Eagle’s medium (BioWhittaker, # BW12-614F) with 10% fetal bovine serum (FBS; Cytiva, #SH30396.03HI), and l-glutamine, and 100 U/ml of penicillin-streptomycin (Gibco, #10378016). Other T-ALL cell lines were cultured in RPMI 1640 supplemented with 10% heat-inactivated FBS, 100 U/ml of penicillin-streptomycin, 2 mM l-glutamine. Surface expression of NTRK1 was assessed in T-ALL cell lines. Cells were stained with APC-conjugated anti-human TrkA (R&D, #FAB175RA) antibody according to the manufacturer’s instructions. Cellular fluorescence data were collected on an LSR II flow cytometer (BD Biosciences) using DIVA software (BD Biosciences). FlowJo v.10.0 (Tree Star) was used for the analysis and visualization.

#### **Viral cloning and transduction**

A gateway-compatible entry clone containing the STAG2 cDNA was obtained from Genecopoeia (#GC-H0274). STAG2 and CRE cDNAs were transferred into a Gateway-compatible Cl20-MSCV-IRES-ametrine lentiviral vector or MSCV-IRES-GFP vector using the LR Clonase II enzyme mix (Life Technologies, #11791100). MiGR1-dE-hNOTCH1-mCherry-Luc was kindly provided from Dr. Ferrando. Constructs were verified by Sanger sequencing. To produce lenti- or retro-virus, 293T cells were cotransfected with pEcopac helper packaging plasmid and (CL20-)MSCV/MiGR1-IRES-GFP/ametrine/mCherry as an empty vector or containing STAG2, dE-hNOTCH1 or CRE by using FuGENE HD Transfection Reagent (Promega, #E2311). Viral supernatants were harvested 48 hours post transfection and filtered through a 0.45 μm filter (Millipore, #SE1M003M00). T-ALL cell lines were infected for 48 hours with LentiBoost (1:100, Sirion Biotech, #NC1889820), Cyclosporin H (8µM, Sigma-Aldrich, #SML1575), and Protamine Sulfate (4 µg/ml) after the over-night pretreatment with Cyclosporin H (8 µM). Transduced ametrine-positive cells were collected by fluorescence-activated cell sorting.

#### **Engineering STAG2 knockout cells**

PF-382-STAG2KO (STAG2 knockout) cells were engineered by CRISPR/Cas9 using Cas9-gRNA ribonucleoprotein (RNP) delivery. Each 3 µl of Cas9 protein (20 µM) and sgRNAs (60 µM) were incubated for 15 minutes at room temperature and mixed with 1 x 10^5^ cells/ml of PF-382 cells in Buffer T (Invitrogen, #MPK1025K). Electroporation was performed using the Neon Transfection System (Thermo Fisher Scientific) with the condition of 1600V, 10ms, and three pulses. After incubating for 72 hours, electroporated cells were sorted into single cells for single cloning. Gene editions were examined by Sanger Sequencing, confirmed loss of STAG2 protein expression by Western blotting, and then performed WGS to validate gene edition and to check off-target effects.

Additional STAG2^-/-^ PF-382 clones were created using CRISPR-Cas9 technology in the Center for Advanced Genome Engineering at St. Jude. Briefly, 1 x 10^6^ PF-382 cells were transiently transfected with precomplexed RNPs consisting of 150 pmol of chemically modified sgRNA (5’ – AAUUCAUUGGCGUGUUAGUA- 3’, Synthego), 50 pmol of Cas9 protein (St. Jude Protein Production Core), and 200 ng of pMaxGFP (Lonza) via nucleofection (Lonza, 4D-Nucleofector™ X-unit) according to the manufacturer’s recommended protocol using solution P3 and program CA-137 in a small (20ul) cuvette. Five days post nucleofection, cells were single-cell sorted by fluorescence-activated cell sorting to enrich for GFP+ cells into 96-well tissue culture treated plates. Cells were clonally expanded and screened for the desired targeted modification via targeted deep sequencing using gene specific primers with partial Illumina adapter overhangs (CAGE1264.STAG2.F – 5’ TGTTCTCTTGCCTACTTTGAGGATT -3’ and CAGE1264.STAG2.R– 5’ GCCAGGGTGCTTGTATGTCG -3’, overhangs not shown) as previously described^27^. NGS analysis of clones was performed using CRIS.py^28^. Final clones were authenticated using the PowerPlex® Fusion System (Promega) performed at the Hartwell Center for Biotechnology at St. Jude Children’s Research Hospital. Final clones tested negative for mycoplasma by the MycoAlertTMPlus Mycoplasma Detection Kit (Lonza).

#### **Western blotting**

Engineered MOLT-14 cells with STAG2 expression add-back and PF-382-STAG2KO were lysed in radioimmunoprecipitation assay buffer supplemented with protease and phosphatase inhibitors (Thermo Fisher Scientific, #1861281). 20 µg of protein of the cell lysate was electrophoresed through 4–12% NuPage Bis-Tris gels (Life Technologies) at 110 V for 130 minutes. Blots were probed with anti-STAG2 (Bethyl, #A302-581A) and anti-β-actin (C4) (Santa Cruz Biotechnology, #sc-47778) antibodies. For imaging, Odyssey DLx (LI-COR) and Image Studio (LI-COR) were used.

#### **Compound screening and cytotoxicity assay**

A total of 2,500 cells were seeded at 30 µl per well in 384-well assay plates (Corning, #8804BC) or 2–4 x 10^5^ cells were seeded at 100 µl per well in 96-well assay plates (Corning, #3603). Assays were performed in triplicate. For compound screening, MOLT-14 and PER-117 cell lines were screened. A total of 2,050 compounds (single point each 10 µM) and 138 compounds (validation, dose-response curve) to be screened were added to assay plates from DMSO stock solutions by acoustic transfer using Echo 650/655 (Beckman coulter). The assay plates were incubated for 72 hours and the CellTiter-Glo (Promega, #G9241) assay was used to determine their sensitivity to each compound. EnVision plate reader (PerkinElmer) was used to measure the luminescence. High-throughput assay data were analyzed using our in-house Robust Interpretation of Screening Experiments (RISE) application written in Pipeline Pilot (Biovia, v17.2.0) according to our previous studies^29,30^. Briefly, with nonlinear regression curve fitting, the area under the drug-response curve per compound is calculated from the area under a fitted hill curve to the data, and a trapezoidal numeric integral of the actual data provided. For cytotoxicity assay, MOLT-14, PER-117, PEER, and LOUCY cell lines were tested with multiple inhibitors including talazoparib (MCE, #HY-16106), C1632 (Cayman, #22401), larotrectinib (LOXO-101, Selleck Chemical LLC, # 50-136-6404), metformin hydrochloride (Millipore, #PHR1084), and vorinostat (Key Organics, #DG-0025) in 9 decreasing concentrations made by serial dilution in 96-well assay plates. DMSO control and blank (medium control) were set appropriately. The assay plates were incubated for 48 hours and cell viability was examined by resazurin cell viability assay (abcam, #ab129732) with Synergy HT plate reader (Bio Tek). GraphPad Prism (version 9) was used to calculate the concentration of drug associated with half-maximal response (EC_50_ values). All assays were performed in triplicate. SynergyFinder 3.0^31^ was used to assess drug synergisms of talazoparib and vorinostat.

PDX drug sensitivity was performed using an assay described previously^32^. Human BM mesenchymal cells immortalized with hTERT (Applied Biological Materials Inc., #T0523) were plated on PhenoPlate 384-well microplates (Revvity, #6057308) at a concentration of 1,500 cells/well and incubated at 37˚ C and 5% CO_2_ for 24 hours in RPMI1640 plus 20% (v/v) fetal bovine serum (Gibco, #10082-147), and 1 µmol/L hydrocortisone (Sigma Aldrich, #H0396). After 24 hours, media is removed from the wells and PDX cells are plated on top of the MSC’s at a concentration of 25,000 cells/well in AIMV serum free media (Gibco, #12055-083) using a VIAFLO384 (Integra). Cells were exposed to a drug library containing 26 compounds (**Appendix Table A19**) in duplicate with DMSO being included as a negative control and maintained at 37˚ C and 5% CO_2_. 96 hours after drug addition, cell viability was measured using CyQUANT Direct Cell Proliferation Assay (Invitrogen, #C35011) and the images were captured using an Operetta (Perkin Elmer) high content imager. Data analysis was performed using the Harmony (version 4.9, Perkin Elmer) software system. For cases in which even the lowest drug concentration killed >50% of leukemia cells, LC_50_ was assigned as half of the minimum tested concentration. Conversely, for cases with >50% viability even at the highest drug concentration, LC_50_ values were assigned as twice of the highest tested concentration. Observed drug LC_50_ values were log-transformed and resulting cell viabilities were used to generate dose response curves using a four-parameter logistic function and to calculate are under the curve (AUC) and half-maximal lethal concentrations (LC_50_) value

### **SUPPLEMENTARY RESULTS**

#### **Genomic subtypes in γδ T-ALL**

The recently described^33^ *LMO2* γδ-like subtype is characterized by a distinct gene expression profile with an expression signature of γδ T-cells with the normal thymocyte/BM single cell mapping, and by detection of γδ TCR rearrangements from WGS/RNAseq data. Such cases exhibit consistent up-regulation of pathways compared to other T-ALL related to immune cell reaction, suggesting the interaction with other immune cells mediated by the γδ T-cell cytokines IL-17 and IFN-γ (**Appendix Fig A3A–F,** and **Appendix Table A9**). Cytologic features include the existence of small blasts with condensed chromatin supporting a relatively mature maturational stage in all analyzed cases (n=3, **Appendix Fig A3G**). This group had distinct but heterogeneous genomic alterations, including *LMO2*-activating alterations, *MYC* translocations, and MYCN P44L mutations^4^ (**Appendix Fig A3H**). Importantly, all *LMO2* γδ-like cases, including those from this study and the reference AALL0434 cohort expressed rearranged γδ TCR on RNAseq analysis (**Appendix Fig A3H**). Furthermore, the *LMO2* γδ-like subtype was not observed in non-γδ T-ALL cases in the St. Jude Total Therapy XV/XVI cohorts^34,35^ indicating it may be specific to γδ T-ALL (**Fig 2B**). For 7 *LMO2* γδ-like cases included in our cohort, three and two patients died due to the transplant toxicity and the primary disease, respectively.

Chr gains with recurrent gains of chromosomes 8, 10, 11, 13q, and 19 were more frequent in γδ T-ALL than in other T-ALL cases (**Fig 2B** and **Appendix Fig A4A**). The timing of the acquisition of aneuploidies was generally synchronous; only one case out of 15 analyzed cases exhibited an asynchronous gain (**Appendix Fig A4B**). All cases harbored either of *PHF6* (68.4%), *MED12* (63.2%), and/or *CTCF* (26.3%) alterations with frequent mutations in JAK-STAT (57.9%) and cell cycle (63.2%) pathway genes (**Appendix Fig A4A**). The gene expression profile of cases with chromosomal gains was similar to that of immature, ETP-like T-ALL (**Fig 2A**). In contrast to B-ALL with high hyperdiploidy, the age at diagnosis in Chr gains T-ALL was older regardless of TCR lineages, and no case diagnosed before age five (**Fig 2D** and **Appendix Fig A4A**).

Other genomic events enriched in γδ T-ALL include rearrangement of *TLX3* and the *PICALM::MLLT10* fusion (**Fig 2B**). The transcriptional profile of the *TLX3*-rearranged γδ cases was intermediate between that of the immature and double positive thymocyte-like (DP-like) TLX3 subtypes (**Fig 2A**), suggestive of developmental origin at the double negative (DN) stage of thymocyte development, where αβ and γδ T-cells bifurcate^36,37^. Consistent with the timing of αβ and γδ bifurcation around DN3a stage,^38^ no alteration was found to activate *BCL11B* (which are too immature), *TLX1*, and *TAL1* (too mature).

#### **TCR repertoire in γδ T-ALL and association with genomic and clinical features**

The T-cell receptor (TCR) rearrangements for the *TRG* and *TRD* loci in γδ T-ALL cases (defined by flow-based expression of γδ TCR) were examined by RNAseq data (or whole genome sequencing [WGS] if no RNAseq data was available) with mixcr^24^ for all 76 cases, and the results were validated using DNA amplicon TRD/G sequencing in 5 cases (**Appendix Table A4**). While Vδ was rearranged in more than half γδ T-ALL as we observe in healthy peripheral blood mononuclear cells repertoire^39^, a variety of Vγ regions were paired with DJ regions except for Vγ6 (**Supplementary Fig S3**). There was no specific usage of TCR repertoire in each genomic subtype, although there were some preferences: Vδ/Vγ4 in *STAG2/LMO2* and *LMO2* γδ-like, and Vδ/Vγ3 in *HOXA9::TCR* (**Supplementary Fig S3B** and **Appendix Table A4**). Though the number was small, all 4 cases with Vδ2/Vγ8 (*STAG2/LMO2* and *TLX3*-rearranged) had poor outcomes due to refractory disease, relapse, or death. However, our TCR analysis is based on RNAseq or WGS not on DNA amplicon sequencing. There are several limitations in RNAseq/WGS-based analysis: incomplete rearrangements may be overlooked; subclonal analysis is challenging due to lower depth than amplicon sequencing; and TCR rearrangements in *TRD* and *TRG* loci were not detected in some γδ T-ALL patients (**Appendix Table A4**). On the other hand, the caution of using DNA amplicon sequencing is that since genomic TCR rearrangements precede their expression, this modality may overestimate the TCR rearrangements of γδ T-ALL patients, who were diagnosed by the expression of γδ TCR by flow cytometry. Therefore, our TCR results and correlation with clinical data should be validated in a larger cohort with consideration of full V-D-J gene sequence and incomplete rearrangements.

#### **Gene-edited model of STAG2 inactivation in T-ALL**

Two gene-edited cell line models: (1) empty vector (EV) or *STAG2* transduced MOLT-14 (*STAG2/LMO2*), and (2) *STAG2* knockout PF382 (*LMO2*-activating mutation; **Fig 4B** and **Appendix Fig A6**) were used to explore the effect of STAG2 inactivation in T-ALL. Restoration of STAG2 expression in MOLT-14 exhibited increased STAG2 binding at intronic and intergenic regions, suggesting STAG2 binding was regained at enhancer regions (**Appendix Fig A6C**). First, H3K27ac binding peaks and corresponding gene expression levels were compared between MOLT14-EV (STAG2 inactivation) and MOLT14-STAG2 (STAG2 restoration). Enforced expression of STAG2 restored two STAG1/STAG2 co-binding sites at the *CD34* locus, resulting in down-regulation of H3K27ac peaks at the *CD34* enhancer and promoter with decreased expression (**Fig 4D** and **Appendix Fig A7**). Pathway analysis using H3K27ac peaks that appeared after STAG2 restoration (those likely lost with STAG2 inactivation) showed enrichment of T-cell differentiation-related pathways (**Appendix Fig A6D**), which included regions of *BCL11B*, *RAG2, EOMES*, and *TBX21*, suggesting that STAG2 inactivation likely drives differentiation arrest.

By using PF382 parental and *STAG2* knockout lines, we further examined the effects of STAG2 inactivation in T-ALL. Polyamine metabolic pathways (**Fig 4F**) were activated by STAG2 inactivation, suggesting that STAG2 inactivation in this context might drive proliferation by polyamine synthesis. Polyamine metabolic pathway is an important metabolic target of MYC,^40^ supporting up-regulated pathways in *STAG2/LMO2* subtype (**Fig 3G**). Similar to the results of STAG2 restoration in the MOLT-14 model, STAG2 inactivation in PF382 cells decreased STAG2 binding at intronic and intergenic regions (**Appendix Fig A6G**).

### **SUPPLEMENTARY FIGURES**

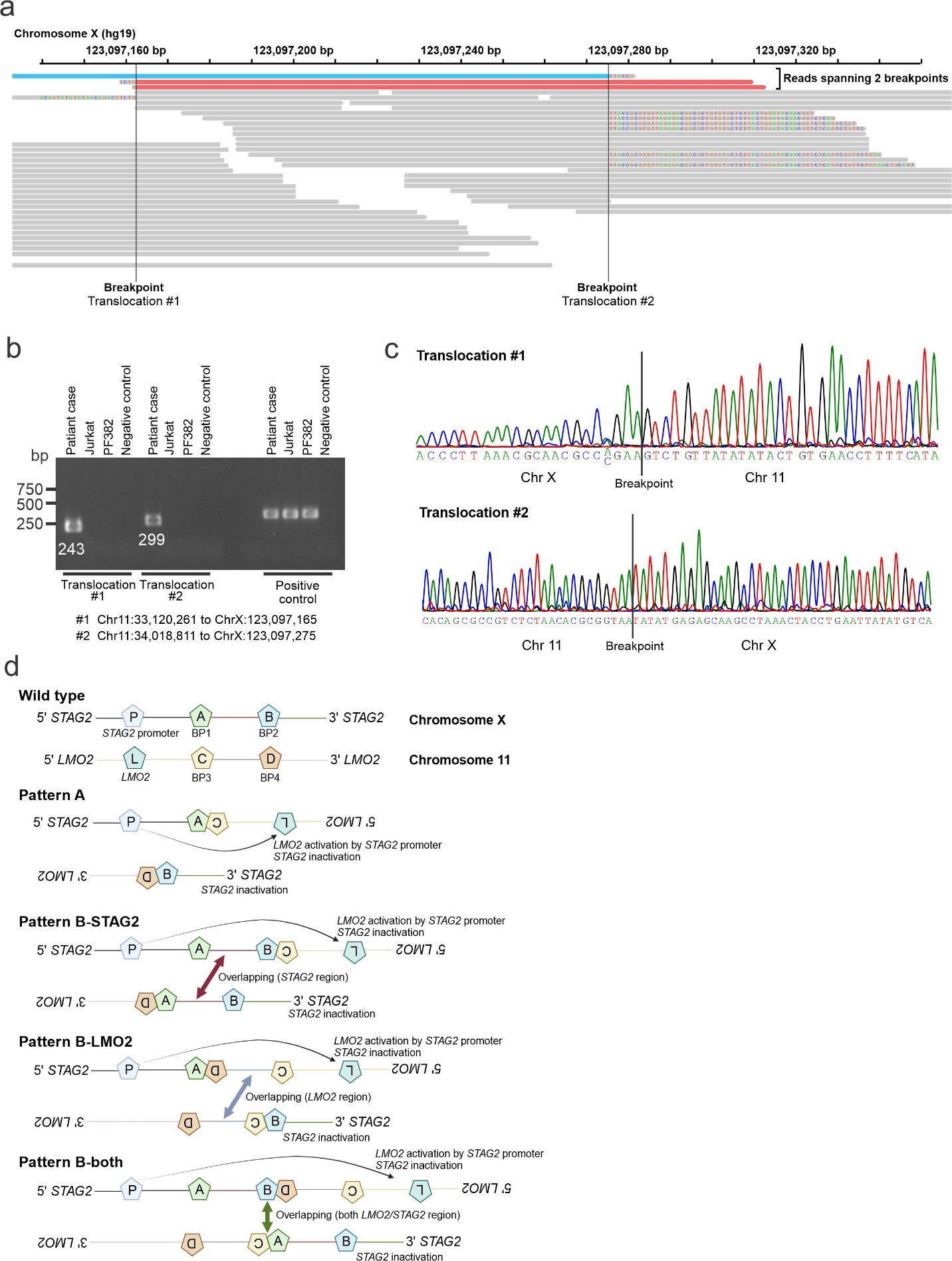

**Supplementary Figure S1. *LMO2::STAG2* translocations.** **a,** The representative example of *LMO2::STAG2* translocations showing multiple breakpoints on chromosome X (*STAG2* intron) in the Integrative Genomics Viewer (IGV). Red and blue colors indicate reads spanning both breakpoints, which are not found on the same read. **b,** Validation of *LMO2::STAG2* translocations by genomic PCR. Two break points were confirmed in the representative case. **c,** Direct sequencing of PCR products. **d,** The patterns of observed *LMO2::STAG2* translocations with multiple breakpoints (BP) in our cohort (4 out of 5 cases), considering allelic usage in *STAG2* and *LMO2*. Pattern A includes reciprocal translocations and may affect one or two alleles. Pattern B contains overlapping regions of *STAG2* and/or *LMO2*, indicating both alleles are affected by translocations.

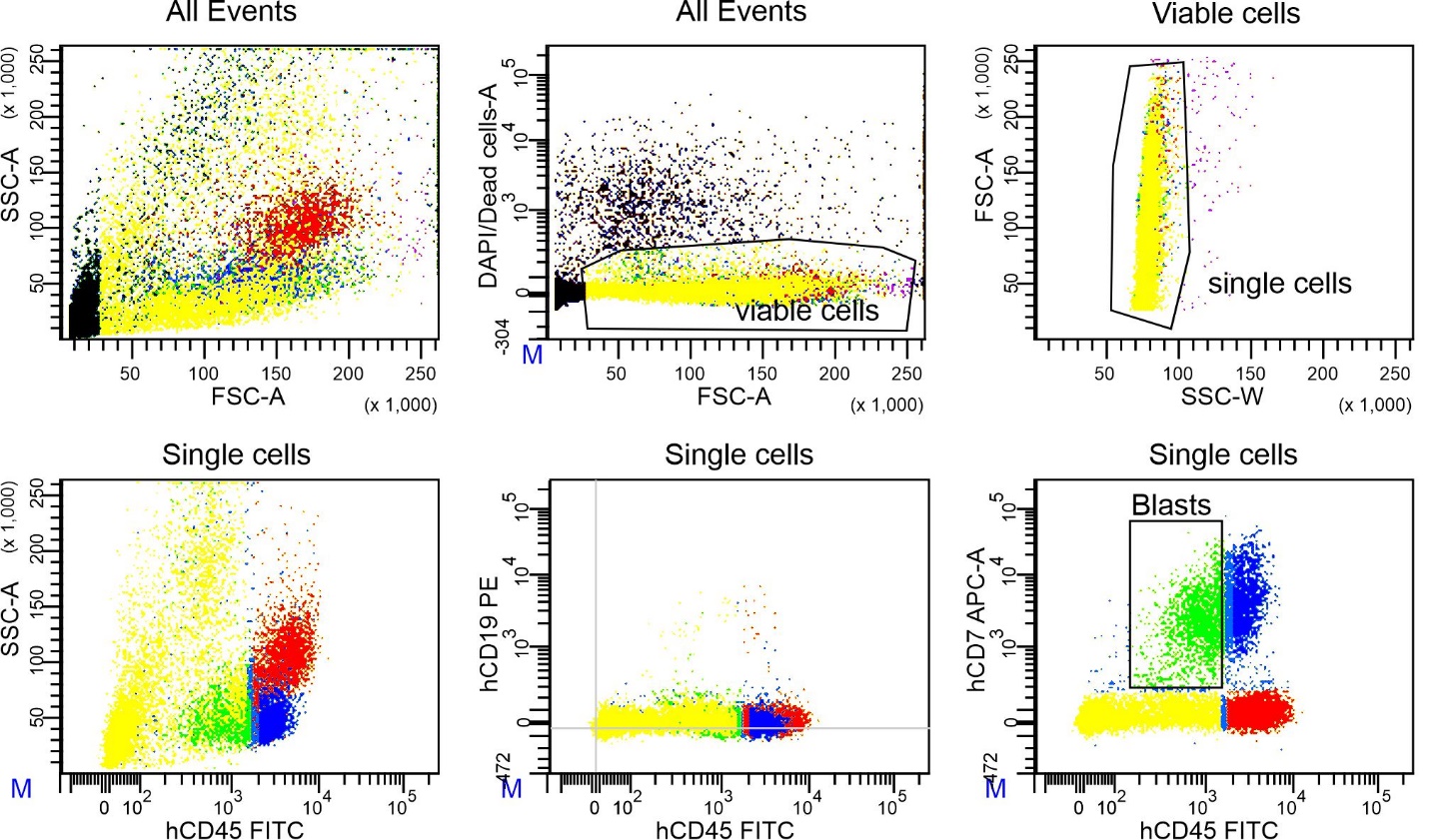

**Supplementary Figure S2. Flow-sorted blast cells (T-ALL).** Leukemic cells of low tumor purity case was sorted based on CD45 dim and CD7 positive population.

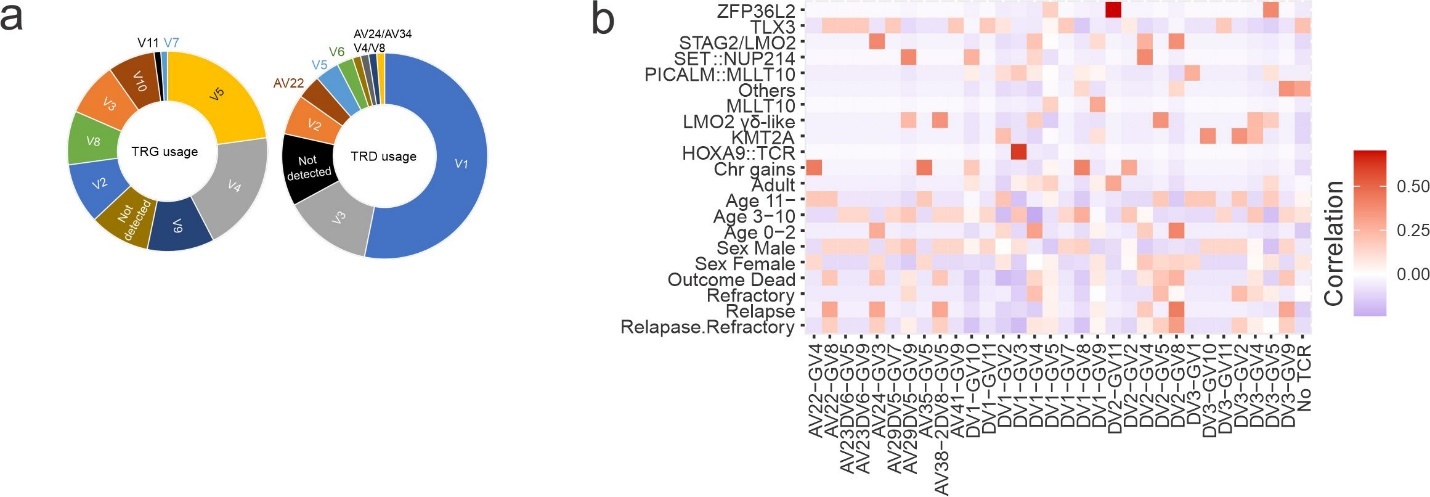

**Supplementary Figure S3. TCR repertoire and clinical and genomic features in γδ T-ALL.** **a,** The V usage in *TRG* and *TRD* loci is shown. While Vδ was rearranged in more than half of γδ T-ALL cases, a variety of Vγ regions except for Vγ6 were used. **b,** The correlation heatmap showing each *TRG*-*TRD* pair of T-cell receptor (TCR) repertoire with genomic subtype, clinical features and outcomes.

### **SUPPLEMENTARY REFERENCES**

1.	Montefiori LE, Bendig S, Gu Z, et al: Enhancer Hijacking Drives Oncogenic BCL11B Expression in Lineage-Ambiguous Stem Cell Leukemia. Cancer Discov 11:2846-2867, 2021

2.	Kimura S, Montefiori L, Iacobucci I, et al: Enhancer retargeting of CDX2 and UBTF::ATXN7L3 define a subtype of high-risk B-progenitor acute lymphoblastic leukemia. Blood 139:3519-3531, 2022

3.	Li H, Durbin R: Fast and accurate short read alignment with Burrows-Wheeler transform. Bioinformatics 25:1754-60, 2009

4.	Liu Y, Easton J, Shao Y, et al: The genomic landscape of pediatric and young adult T-lineage acute lymphoblastic leukemia. Nat Genet 49:1211-1218, 2017

5.	Rausch T, Zichner T, Schlattl A, et al: DELLY: structural variant discovery by integrated paired-end and split-read analysis. Bioinformatics 28:i333-i339, 2012

6.	Chen X, Schulz-Trieglaff O, Shaw R, et al: Manta: rapid detection of structural variants and indels for germline and cancer sequencing applications. Bioinformatics 32:1220-2, 2016

7.	Robinson JT, Thorvaldsdottir H, Winckler W, et al: Integrative genomics viewer. Nat Biotechnol 29:24-6, 2011

8.	Chen X, Gupta P, Wang J, et al: CONSERTING: integrating copy-number analysis with structural-variation detection. Nat Methods 12:527-30, 2015

9.	Klambauer G, Schwarzbauer K, Mayr A, et al: cn.MOPS: mixture of Poissons for discovering copy number variations in next-generation sequencing data with a low false discovery rate. Nucleic Acids Res 40:e69, 2012

10.	Brady SW, Roberts KG, Gu Z, et al: The genomic landscape of pediatric acute lymphoblastic leukemia. Nat Genet 54:1376-1389, 2022

11.	Gu Z, Churchman ML, Roberts KG, et al: PAX5-driven subtypes of B-progenitor acute lymphoblastic leukemia. Nat Genet 51:296-307, 2019

12.	Seki M, Kimura S, Isobe T, et al: Recurrent SPI1 (PU.1) fusions in high-risk pediatric T cell acute lymphoblastic leukemia. Nat Genet 49:1274-1281, 2017

13.	Dobin A, Davis CA, Schlesinger F, et al: STAR: ultrafast universal RNA-seq aligner. Bioinformatics 29:15-21, 2013

14.	Anders S, Pyl PT, Huber W: HTSeq--a Python framework to work with high-throughput sequencing data. Bioinformatics 31:166-9, 2015

15.	Leek JT, Johnson WE, Parker HS, et al: The sva package for removing batch effects and other unwanted variation in high-throughput experiments. Bioinformatics 28:882-3, 2012

16.	Love MI, Huber W, Anders S: Moderated estimation of fold change and dispersion for RNA-seq data with DESeq2. Genome Biol 15:550, 2014

17.	Wu T, Hu E, Xu S, et al: clusterProfiler 4.0: A universal enrichment tool for interpreting omics data. Innovation (Camb) 2:100141, 2021

18.	Barinka J, Hu Z, Wang L, et al: RNAseqCNV: analysis of large-scale copy number variations from RNA-seq data. Leukemia 36:1492-1498, 2022

19.	Mumbach MR, Rubin AJ, Flynn RA, et al: HiChIP: efficient and sensitive analysis of protein-directed genome architecture. Nat Methods 13:919-922, 2016

20.	Juric I, Yu M, Abnousi A, et al: MAPS: Model-based analysis of long-range chromatin interactions from PLAC-seq and HiChIP experiments. PLoS Comput Biol 15:e1006982, 2019

21.	Dickerson KM, Qu C, Gao Q, et al: ZNF384 Fusion Oncoproteins Drive Lineage Aberrancy in Acute Leukemia. Blood Cancer Discov 3:240-263, 2022

22.	Zhang Y, Liu T, Meyer CA, et al: Model-based analysis of ChIP-Seq (MACS). Genome Biol 9:R137, 2008

23.	McLean CY, Bristor D, Hiller M, et al: GREAT improves functional interpretation of cis-regulatory regions. Nat Biotechnol 28:495-501, 2010

24.	Bolotin DA, Poslavsky S, Mitrophanov I, et al: MiXCR: software for comprehensive adaptive immunity profiling. Nat Methods 12:380-1, 2015

25.	Bruggemann M, Kotrova M, Knecht H, et al: Standardized next-generation sequencing of immunoglobulin and T-cell receptor gene recombinations for MRD marker identification in acute lymphoblastic leukaemia; a EuroClonality-NGS validation study. Leukemia 33:2241-2253, 2019

26.	Bystry V, Reigl T, Krejci A, et al: ARResT/Interrogate: an interactive immunoprofiler for IG/TR NGS data. Bioinformatics 33:435-437, 2017

27.	Narina S, Connelly JP, Pruett-Miller SM: High-Throughput Analysis of CRISPR-Cas9 Editing Outcomes in Cell and Animal Models Using CRIS.py. Methods Mol Biol 2631:155-182, 2023

28.	Connelly JP, Pruett-Miller SM: CRIS.py: A Versatile and High-throughput Analysis Program for CRISPR-based Genome Editing. Sci Rep 9:4194, 2019

29.	Chang Y, Min J, Jarusiewicz JA, et al: Degradation of Janus kinases in CRLF2-rearranged acute lymphoblastic leukemia. Blood 138:2313-2326, 2021

30.	Chang Y, Keramatnia F, Ghate PS, et al: The orally bioavailable GSPT1/2 degrader SJ6986 exhibits in vivo efficacy in acute lymphoblastic leukemia. Blood 142:629-642, 2023

31.	Ianevski A, Giri AK, Aittokallio T: SynergyFinder 3.0: an interactive analysis and consensus interpretation of multi-drug synergies across multiple samples. Nucleic Acids Res 50:W739-W743, 2022

32.	Rowland L, Smart B, Brown A, et al: Ex vivo Drug Sensitivity Imaging-based Platform for Primary Acute Lymphoblastic Leukemia Cells. Bio Protoc 13:e4731, 2023

33.	Pölönen P, Elsayed A, Di Giacomo D, et al: Comprehensive Genome Characterization of Childhood T-ALL Links Oncogene Activation Mechanism and Subtypes to Prognosis. Blood 140:1727-1729, 2022

34.	Pui CH, Campana D, Pei D, et al: Treating childhood acute lymphoblastic leukemia without cranial irradiation. N Engl J Med 360:2730-41, 2009

35.	Jeha S, Pei D, Choi J, et al: Improved CNS Control of Childhood Acute Lymphoblastic Leukemia Without Cranial Irradiation: St Jude Total Therapy Study 16. J Clin Oncol 37:3377-3391, 2019

36.	Yui MA, Rothenberg EV: Developmental gene networks: a triathlon on the course to T cell identity. Nat Rev Immunol 14:529-45, 2014

37.	Roels J, Kuchmiy A, De Decker M, et al: Distinct and temporary-restricted epigenetic mechanisms regulate human alphabeta and gammadelta T cell development. Nat Immunol 21:1280-1292, 2020

38.	Ciofani M, Zuniga-Pflucker JC: Determining gammadelta versus alphass T cell development. Nat Rev Immunol 10:657-63, 2010

39.	Kallemeijn MJ, Kavelaars FG, van der Klift MY, et al: Next-Generation Sequencing Analysis of the Human TCRgammadelta+ T-Cell Repertoire Reveals Shifts in Vgamma- and Vdelta-Usage in Memory Populations upon Aging. Front Immunol 9:448, 2018

40.	Casero RA, Jr., Murray Stewart T, Pegg AE: Polyamine metabolism and cancer: treatments, challenges and opportunities. Nat Rev Cancer 18:681-695, 2018
